# Noninvasive diagnosis of secondary infections in COVID-19 by sequencing of plasma microbial cell-free DNA

**DOI:** 10.1101/2022.09.09.22279790

**Authors:** Grace Lisius, Radha Duttagupta, Asim A. Ahmed, Matthew Hensley, Nameer Al-Yousif, Michael Lu, William Bain, Faraaz Shah, Caitlin Schaefer, Shulin Qin, Xiaohong Wang, Yingze Zhang, Kevin J. Mitchell, Ellen K. Hughes, Jana L. Jacobs, Asma Naqvi, Ghady Haidar, John W. Mellors, Barbara Methé, Bryan J. McVerry, Alison Morris, Georgios D. Kitsios

**Author notes:** Corresponding Author: Georgios D. Kitsios, MD, PhD, Assistant Professor of Medicine, Division of Pulmonary, Allergy and Critical Care Medicine, University of Pittsburgh Medical Center, Address: UPMC Montefiore Hospital, NW628, 3459 Fifth Avenue, Pittsburgh, PA 15213. **Funding information:** Dr. Kitsios: University of Pittsburgh Clinical and Translational Science Institute, COVID-19 Pilot Award; NIH (K23 HL139987; R03 HL162655). Dr. Bain: Career Development award number IK2 BX004886 from the U.S. Department of Veterans Affairs Biomedical Laboratory R&D (BLRD) Service. Caitlin Schaefer: NIH (P01 HL114453).

## Abstract

**Background:** Secondary infection (SI) diagnosis in COVID-19 is challenging, due to overlapping clinical presentations, practical limitations in obtaining samples from the lower respiratory tract (LRT), and low sensitivity of microbiologic cultures.

**Research Question:** Can metagenomic sequencing of plasma microbial cell-free DNA (mcfDNA-Seq) help diagnose SIs complicating COVID-19?

**Study Design and Methods:** We enrolled 42 inpatients with COVID-19 classified as microbiologically-confirmed SI (Micro-SI, n=8), clinically-diagnosed SI (Clinical-SI, n=13, i.e. empiric antimicrobials), or no clinical suspicion for SI (No-Suspected-SI, n=21) at time of enrollment. From baseline and follow-up plasma samples (days 5 and 10 post-enrollment), we quantified mcfDNA for all detected microbes by mcfDNA sequencing and measured nine host-response biomarkers. From LRT samples among intubated subjects, we quantified bacterial burden with 16S rRNA gene quantitative PCR.

**Results:** We performed mcfDNA-Seq in 82 plasma samples. Sequencing was successful in 60/82 (73.2%) samples, which had significantly lower levels of human cfDNA than failed samples (p<0.0001). McfDNA detection was significantly higher in Micro-SI (15/16 [94%]) compared to Clinical-SI samples (8/14 [57%], p=0.03), and unexpectedly common in No-Suspected-SI samples (25/30 [83%]), similar to detection rate in Micro-SI. We detected culture-concordant mcfDNA species in 13/16 Micro-SI samples (81%) and mcfDNA levels tracked with SI outcome (resolution or persistence) under antibiotic therapy. McfDNA levels correlated significantly with LRT bacterial burden (r=0.74, p=0.02) as well as plasma biomarkers of host response (white blood cell count, IL-6, IL-8, and SPD, all p<0.05). Baseline mcfDNA levels were predictive of worse 90-day survival (hazard ratio 1.30 [1.02-1.64] for each log_10_ mcfDNA, p=0.03).

**Interpretation:** High circulating levels of mcfDNA in a substantial proportion of patients with COVID-19 without clinical suspicion for SI suggest that SIs may often remain undiagnosed. McfDNA-Seq, when clinically available, can offer a non-invasive diagnostic tool for pathogen identification, with prognostic value on host inflammatory response and clinical outcomes.

## Introduction

Secondary bacterial or fungal infections complicate the course of up to 50% of hospitalized patients with COVID-19 and contribute to worse clinical outcomes.^1–3^ Diagnosis of secondary infections (SI) has presented clinicians with major challenges during the pandemic. Readily available indicators of clinical infection such as fever, leukocytosis or consolidations on chest imaging cannot distinguish isolated SARS-CoV-2 infection from a secondary pneumonia by super-infecting organisms.^4,5^ Blood cultures are obtained routinely when a SI is suspected in COVID-19, but their diagnostic yield is low, especially in the case of secondary pneumonias, when direct lower respiratory tract (LRT) sampling is recommended.^6^ LRT sampling for microbiologic studies can be challenging, such as in severely hypoxemic patients on non-invasive respiratory support, and can have low diagnostic sensitivity, due to antecedent antibiotics or slow/fastidious organism growth, as in the case of fungal pathogens.^7^

The challenges of SI diagnostic work-up and variable clinical practices have hindered acquisition of accurate SI incidence estimates in severely ill patients with COVID-19. With the prevailing diagnostic uncertainty, empiric antibiotics have been prescribed in 75-85% of hospitalized patients, often initiated due to clinical deterioration and then empirically continued, even in the absence of diagnostic evidence supporting an SI.^8,9^ Overcoming the diagnostic challenges of SI with reliable, sensitive and non-invasive techniques could optimize diagnostic yield, enabling antibiotic targeting and stewardship.^4^

Metagenomic sequencing of plasma microbial cell-free DNA (mcfDNA) can offer a non-invasive, sensitive diagnostic tool for SIs caused by DNA pathogens. Plasma metagenomics in sepsis provides rapid and actionable results, with 95% concordance with blood-cultured organisms, facilitating adjustments of empiric antimicrobials to targeted therapy.^10^ We have previously provided proof-of-concept evidence that patients with COVID-19 with high levels of circulating mcfDNA of common respiratory pathogens were at risk of worse clinical outcome.^11^ Although the ability to comprehensively assess for SI-causal pathogens with non-invasive blood samples is appealing, feasibility and clinical validity data for mcfDNA sequencing in COVID-19 are needed. In this study, we report the analyses of a cohort of 42 hospitalized patients with severe COVID-19 who were comprehensively screened for SI pathogens with serial plasma mcfDNA sequencing.

## Methods

### Clinical cohort enrollment and COVID-19 diagnosis

From April 2020 through September 2020, we conducted an observational, prospective cohort study of hospitalized patients with acute hypoxemic respiratory failure and confirmed severe COVID-19 pneumonia as previously described.^12,13^ The University of Pittsburgh Institutional Review Board (IRB) approved the protocols and we obtained written or electronic informed consent by all participants or their surrogates in accordance with the Declaration of Helsinki. We diagnosed COVID-19 based on institutional clinical criteria (clinical symptoms, hypoxemia and abnormal chest radiographic findings) with confirmatory molecular testing (positive SARS-CoV-2 nasopharyngeal quantitative polymerase chain reaction [qPCR]).

### Biospecimen collection and experiments

We collected blood samples in EDTA tubes on enrollment (baseline - day 1) for centrifugation, separation and storage of plasma and additional blood constituents until conduct of experiments. We also collected repeat blood samples on days 5 and 10 post-enrollment from critically ill patients who remained hospitalized in the intensive care unit (ICU). We collected endotracheal aspirates (ETA) from mechanically-ventilated patients concurrently with blood sample acquisition.

We conducted plasma microbial cell-free DNA metagenomic sequencing (mcfDNA-Seq) with the Karius Test^®^ [Karius Inc., Redwood City, CA] and classified the derived metagenomic sequences as human (hcfDNA) vs. microbial (mcfDNA). Based on minimum sequencing coverage metric required for quality control, we classified sequencing runs as “Pass”, “Qualitatively Pass” or “Failed”. All microbes were reported at species level and included a quantitative measure of abundance expressed as DNA molecules per microliter (MPMs), with the exception of the “Qualitative Pass” calls where the reported organism could not be quantified. We classified microbes identified by mcfDNA-Seq into recognized respiratory pathogens vs. microbes of unclear clinical importance.^14^ For contextualization, we compared mcfDNA levels among subjects with COVID-19 with our previously published dataset of mechanically ventilated patients with and without pneumonia.^14^

To profile the host response, we measured plasma levels of nine prognostic biomarkers (eight interleukin [IL]-6, IL-8, pentraxin-3, procalcitonin, receptor for advanced glycation end products [RAGE], angiopoietin-2 (Ang-2), suppression of tumorigenicity [ST]-2, and tumor necrosis factor receptor [TNFR]-1 with Luminex, and surfactant Protein D [SPD] with ELISA; R&D Systems, Minnesota). We measured plasma SARS-CoV-2 viral levels (vRNA) following RNA extraction and qPCR amplification with a sensitive in-house method.^15^ From available ETA samples, we separately extracted genomic DNA and RNA and performed bacterial 16S rRNA gene and SARS-CoV-2 RNA qPCR for assessment of bacterial and viral load in the LRT, respectively.^5,16^

### Diagnosis and classification of secondary infections

We reviewed all available clinical, microbiologic and antimicrobial treatment data around the timing of baseline samples (+/-3 days) for patients with COVID-19 to ascertain the presence/absence or clinical suspicion for SI. We classified subjects in three groups, assessed by two reviewers:

a. Microbiologically-Confirmed Secondary Infection (**Micro-SI**), when typical pathogenic microbes were isolated on clinical biospecimen cultures (blood, LRT, urine or tissue) and the subject was receiving antimicrobials for the documented infection.
b. Clinically-Diagnosed Secondary Infections (**Clinical-SI**), when empiric antimicrobials were administered without microbiologic SI confirmation.
c. “No Clinical Suspicion for SI” (**No-Suspected-SI**), when microbiologic workup for SI was negative or not performed, and no empiric antimicrobials were prescribed.

We repeated classifications into the Micro-SI, Clinical-SI and No-Suspected-SI categories at the timing of follow-up samples (days 5 and 10 post-enrollment) from updated clinical and microbiologic data, when available. We quantified radiographic edema of baseline chest x-rays with the Radiographic Assessment of Lung Edema (RALE) score with the use of the *PulmAnnotator* software.^17,18^

### Statistical analysis

For “Pass” samples with zero microbial calls, we assigned a microbial MPM of “1” to allow for log_10_-transformations, and utilized log_10_-transformed values of plasma biomarkers and mcfDNA levels for analyses due to non-normal data. We compared clinical variables, biomarker and cfDNA levels between different categories (i.e. sequencing run success, SI clinical groups) and the historical non-COVID-19 cohort using non-parametric tests, Wilcoxon signed-rank and Fischer’s exact tests. We examined for correlations between biomarkers and cfDNA levels with the Pearson’s method, adjusted for multiple comparisons with the Benjamini-Hochberg method. We analyzed the impact of baseline mcfDNA levels on 90-day survival by Kaplan-Meier curves stratified by total mcfDNA tertiles and by constructing Cox proportional age-adjusted hazards models. We performed all analyses with the R software (R version 3.5.1).

## Results

### Clinical cohort characteristics

We included 42 patients (median age 65.1 years, 62% men), who contributed a total of 82 plasma samples for cfDNA sequencing (median of 2 samples per subject). From available baseline data, we classified subjects as Micro-SI (n=8), Clinical-SI (n=13) and No-Suspected-SI (n=21) (Table 1). We found lower rates of invasive mechanical ventilation (p = 0.05) and lower total white blood cell count (p=0.04) in Clinical-SI patients and no other significant clinical variable differences between SI groups. Plasma vRNA was notably lower in Micro-SI (p≤0.05). In biomarker comparisons, we found significant between-group differences in IL-8 and Ang-2 levels (lower in Clinical-SI).

**TABLE 1:**
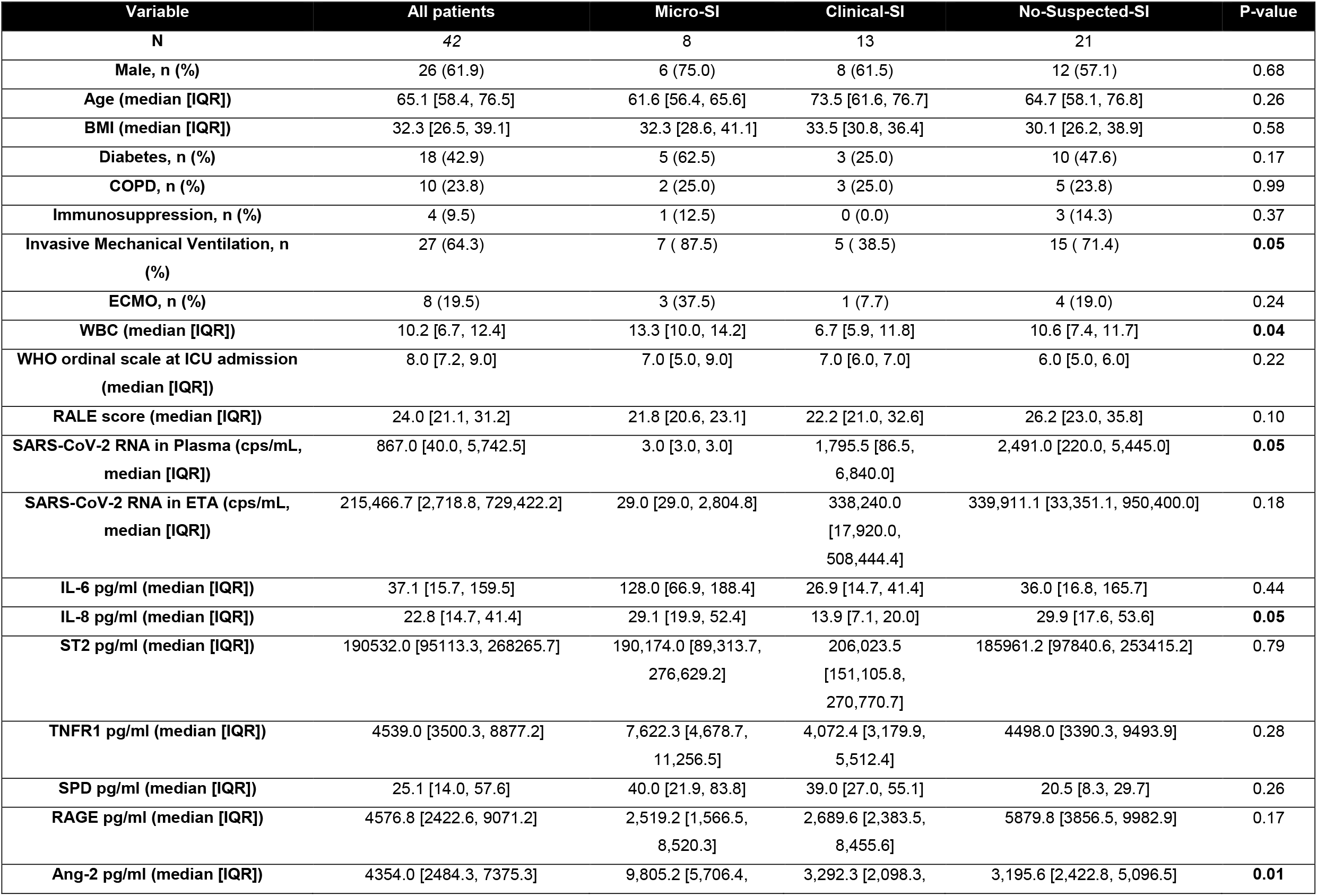
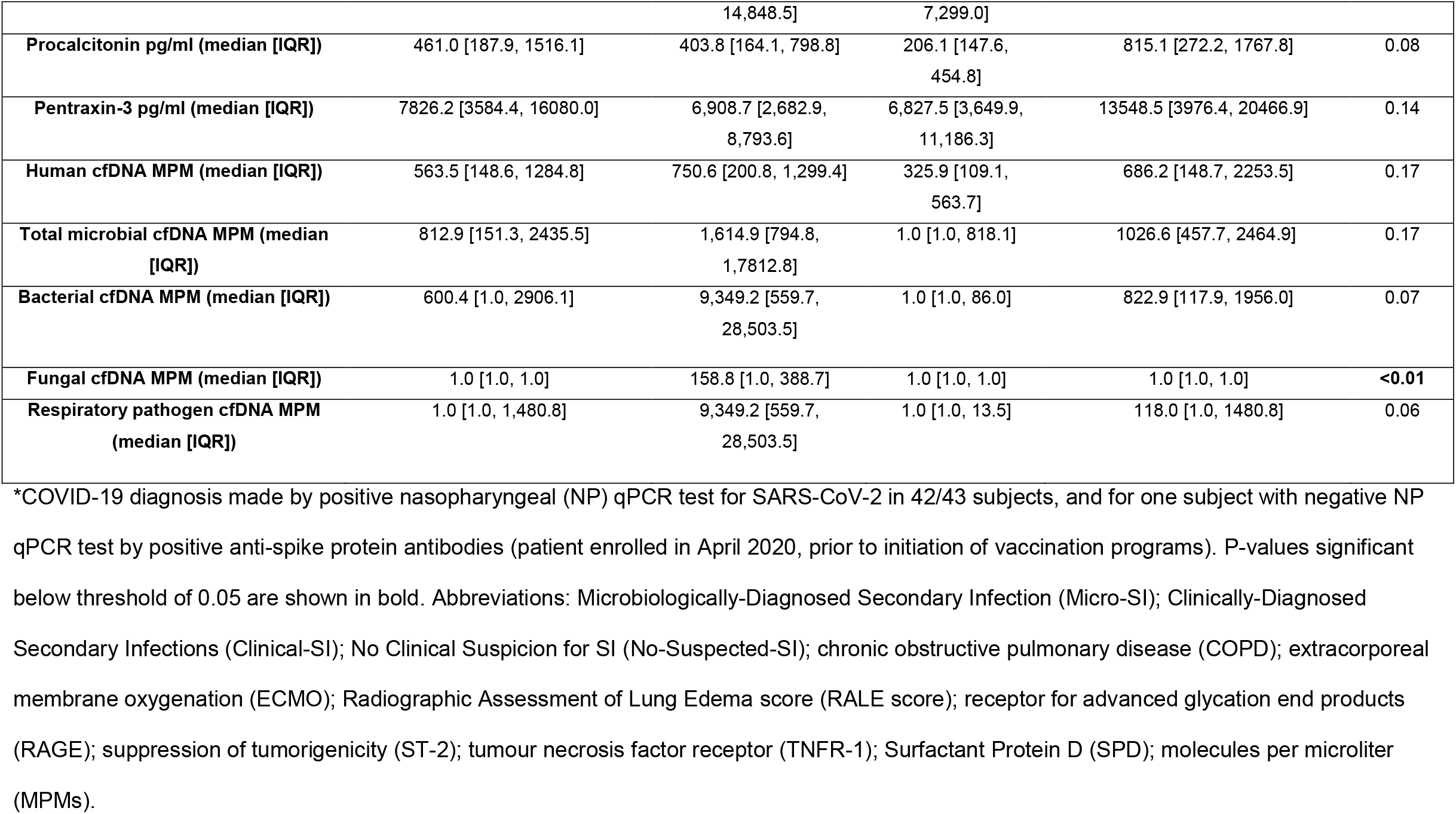
Cohort characteristics by COVID-19 and SI category.

### Outputs of plasma metagenomic sequencing runs

Based on the sequencing run success criteria of the 82 samples, we classified 52 (63%) as “Pass” (successful), eight (10%) as “Qualitative Pass” and 22 samples (27%) as “Fail”. “Pass” samples had significantly lower levels of hcfDNA compared to “Qualitative Pass”, or “Fail” samples (p<0.0001, e-Figure 1). We also found that a successful sequencing run (“Pass”) on a Day 1 sample for a given subject was significantly associated with a “Pass” run on a Day 5 sample (Fisher’s odds ratio 11.9, 95% confidence interval-CI [1.00-703.8], p=0.04). From “Qualitative Pass” samples, we utilized only qualitative information about identified microbial species, whereas we utilized quantitative mcfDNA data (expressed as MPMs) from “Pass” samples only.

### Plasma metagenomics results by SI category

Among the 53 “Pass” samples across all time-points, Micro-SI diagnosis had more samples positive for mcfDNA (i.e. DNA reads from at least one microbial species reported, 15/16 [94%] samples) compared to Clinical-SI (8/14 [57%], p=0.03). Unexpectedly, a high proportion of No-Suspected-SI samples were positive for mcfDNA (25/30 [83%]), similar to Micro-SI samples. All eight “Qualitative Pass” samples, one Clinical-SI and seven No-Suspected-SI, were positive for mcfDNA, with 3/8 samples reporting ≥2 microbial species.

Among baseline “Pass” samples (n = 25), 18 (72%) were positive for mcfDNA, with a median (interquartile range – IQR) of 812.9 (151.3-2435.5) MPMs per sample, 59.1% of which corresponded to typical pathogenic organisms. Stratified by SI categories, 4/5 (80.0%) Micro-SI, 3/8 (37.5%) Clinical-SI and 11/12 (91.6%) No-Suspected-SI samples were positive for mcfDNA, with a statistically significant higher proportion of positive samples in No-Suspected-SI compared to Clinical-SI samples (Fisher’s p=0.01). We found no significant differences in hcfDNA, total mcfDNA and pathogen mcfDNA MPMs between SI groups (Figure 1A-C), although Micro-SI samples had numerically higher total and pathogen mcfDNA MPM levels (Table 1).

**Figure 1:**
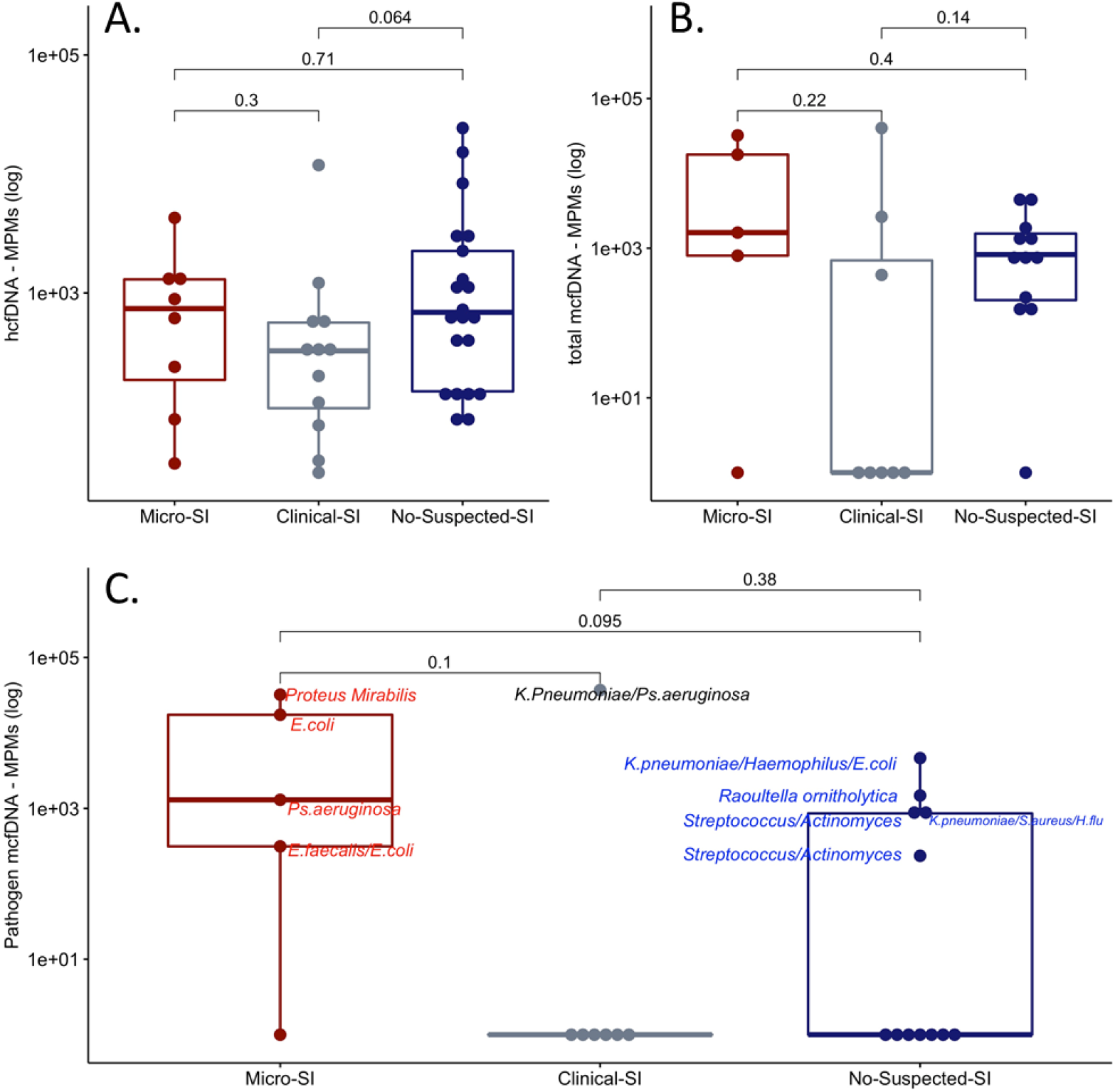
Clinical classification of secondary infection diagnosis among patients with COVID-19 did not show significant differences in human (A) or microbial cell-free DNA levels (B-C). Subjects classified as Microbiologically-Confirmed Secondary Infection (Micro-SI) had numerically higher but statistically non-significant different levels of total and pathogen microbial cell-free DNA (mcfDNA) compared to subjects classified as Clinically-Diagnosed Secondary Infections (Clinical-SI) or those with no Clinical Suspicion for SI (No-Suspected-SI). Pairwise comparisons were conducted with Wilcoxon test. In panel C, the most abundant microbial species in each of the positive samples for pathogen mcfDNA are shown.

Among baseline “Pass” samples in Micro-SI subjects, plasma metagenomics reported mcfDNA from the culprit organisms identified by cultures in 3/5 (60.0%) cases (*E*.*coli* in subject 4, *Pseudomonas aeruginosa* in subject 3 and *Proteus mirabilis* in subject 6) (e-Figure 2, eTable 2). The two discordant Micro-SI samples involved two probable ventilator-associated pneumonia (VAP) cases: subject 1 with negative mcfDNA-Seq when LRT cultures taken three days prior to plasma sampling showed rare *E*.*coli* growth, and case subject 2 with light Methicillin-sensitive *S. aureus* growth in LRT cultures taken two days prior to plasma sampling was deemed as culprit, but mcfDNA-Seq reported *E. faecalis* and *E*.*coli* mcfDNA. When considering all available Micro-SI samples (five baseline and 11 follow-up samples), mcfDNA-Seq results were concordant with cultures in 13/16 (81.3%) of comparisons.

Baseline samples from Clinical-SI subjects contained mcfDNA in 37.5 % of cases (3/8). Case 11 had high mcfDNA levels for plausible pathogens (*K. pneumoniae* and *P. aeruginosa*; Figure 1C, e-Figure 2, and eTable 2), which had not been detected by blood cultures (no LRT specimen cultures available). For the other two Clinical-SI subjects (15 and 21), reported mcfDNA belonged mostly to Human Herperviruses, which could represent viral re-activation in context of critical illness.^19^

Among No-Suspected-SI patients at baseline, 11/12 (91.6%) samples were positive for mcfDNA, with five samples showing >100 pathogen mcfDNA MPMs in the range of Micro-SI subjects (annotated species in Figure 1C, e-Figure 2). The remaining six subjects with mcfDNA revealed organisms of unclear clinical significance: Herpesvirus DNA in four subjects (27, 28, 38 and 42), and gram-positive bacteria often considered as skin contaminants in blood cultures (*Lactobacillus gasseri* and *S. epidermidis*) in three subjects (38, 41 and 42). Notably, 3/4 (75%) of the baseline “Qualitative Pass” samples for No-Suspected-SI subjects (25, 30, 36) were also positive for pathogen mcfDNA, even though the absolute burden of mcfDNA MPMs could not be reliably estimated.

### Comparison of circulating cfDNA burden in COVID-19 vs. historic non-COVID samples

Comparing against published data from our group for mechanically ventilated patients with microbiologically confirmed pneumonia (n=26), clinically diagnosed pneumonia (n=41) and uninfected controls (n=16, intubated for airway protection or due to cardiogenic pulmonary edema), we found markedly higher levels of hcfDNA in subjects with COVID-19 compared to all non-COVID patient groups (e-Figure 3A, p<0.005 for all comparisons). Patients without COVID with microbiologically confirmed pneumonia had higher mcfDNA levels compared to No-Suspected-SI in patients with COVID-19, who in turn had markedly higher mcfDNA levels compared to uninfected controls (e-Figure 3B-C, all p<0.05).

### Trajectories of plasma cfDNA levels in subjects with COVID-19

We updated SI classifications at the time of the follow-up sample acquisition and found that few subjects transitioned into a different SI diagnostic category (4/26 [15%] and 4/16 [25%] for days 5 and 10, respectively, e-Figure 4, 5). Micro-SI subjects had significantly higher total and pathogen mcfDNA levels compared to No-Suspected-SI subjects at post-enrollment day 5 (p<0.05, e-Figure 5).

We examined subject-level trajectories in four cases with persistent culture-proven SI or persistent pathogen mcfDNA detection on available longitudinal samples (Figure 2). Subject 3 was diagnosed with resistant *Pseudomonas aeruginosa* VAP with clinical and microbiologic relapse, demonstrating persistently elevated *Pseudomonas aeruginosa* mcfDNA for >30 days on repeated sampling (Figure 2). For subject 4 McfDNA-Seq demonstrated a transition in detected bacterial species in follow-up samples corresponding to two different VAP episodes (by *E. coli* and *Pseudomonas aeruginosa* respectively). Subject 6 had unexplained, persistent *Proteus mirabilis* mcfDNA detection following treatment of a urinary tract infection, but was found to have a late parotid gland abscess that was drained and cultures grew *Proteus mirabilis*. Last, for subject 26 diagnosed as No-Suspected-SI (i.e. no empiric antimicrobials) who died on day 6 with multi-organ failure attributed to isolated SARS-CoV-2 infection, we found high pathogen mcfDNA levels, including *Klebsiella pneumoniae*, raising concern for the possibility of an undetected and undertreated SI.

**Figure 2:**
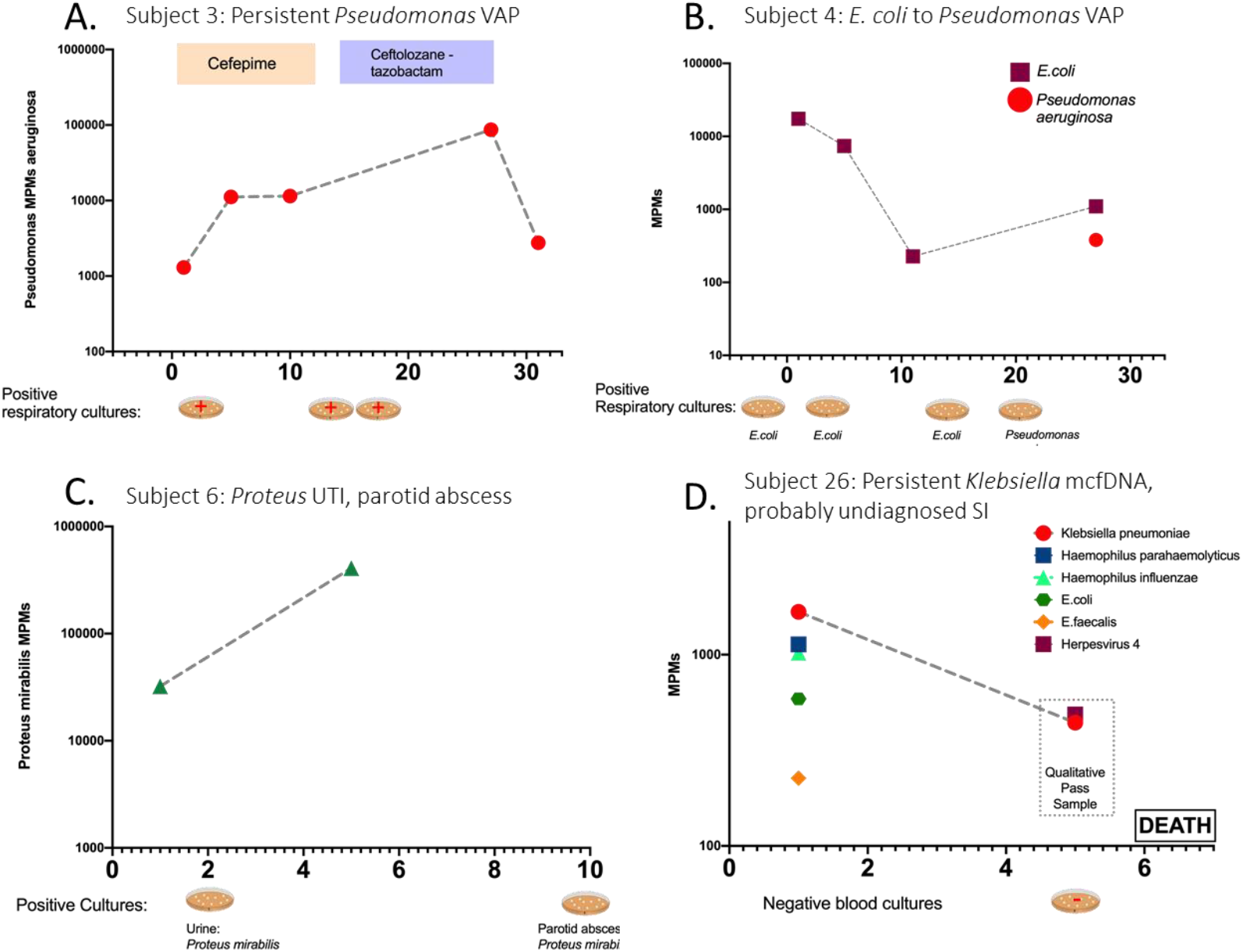
Comparison of plasma mcfDNA sequencing with microbiologic/clinical diagnoses across the timeline of four subjects with serial sampling. Species-specific microbial MPMs (y-axis) are shown across sequential sampling days from enrollment, (x-axis). Petri dish graphics along the x-axis denote the timing and results of clinically obtained microbiological testing. Subject 3 (A) was an immunocompetent patient with a persistent culture-confirmed *Pseudomonas* VAP that persisted through the antimicrobials, with timing and therapy noted above. Subject 3 had persistently elevated *Pseudomonas* mcfDNA levels throughout the extended infection course, which ultimately correlated with clinical improvement (A). Subject 4 (B) had sequential VAPs with changing pathogens detected on invasive respiratory cultures, *E. coli* followed by *Pseudomonas*, which was concordantly reported on noninvasive mcfDNA sampling. Subject 6 (C) was a patient with diabetes who was initially found to have a resistant *Proteus* urinary infection, and a subsequent persistent septic clinical picture, later found to have a polymicrobial, including *Proteus*, parotid gland abscess. *Proteus* mcfDNA levels remained elevated in Subject 6 during initial antimicrobial therapy for urinary infection, suggesting the persistent source of infection. Subject 26 (D) was clinically determined No-Suspicion-for-SI, but had a deteriorating clinical course, without cultures obtained for 5 days or empiric antimicrobials, and ultimately died of shock and multisystem organ failure. Noninvasive testing revealed persistent levels of *Klebsiella pneumonia* mcfDNA levels, which suggests an undiagnosed SI may have contributed to the clinical course.

### McfDNA is associated with plasma host-response biomarkers and LRT bacterial burden

We next examined the relationship between mcfDNA-Seq results and clinical/laboratory parameters of COVID-19 severity. We found no significant correlation between mcfDNA and SARS-CoV-2 viral load (plasma and ETA vRNA), radiographic severity (quantified by RALE scores) or clinical severity by WHO ordinal scale. However, there was a weak, significant correlation between hcfDNA and RALE scores (r=0.33, p=0.03).

We then examined for correlations between cfDNA levels and host-response biomarkers measured in plasma. Total and pathogen mcfDNA levels were significantly correlated with IL-6, IL-8 and SPD levels, whereas hcfDNA levels were significantly correlated with IL-6, IL-8, pentraxin-3 and procalcitonin levels following adjustment for multiple comparisons (Figure 3). Pathogen mcfDNA levels were significantly correlated with 16S rRNA gene copies by qPCR in ETA specimens, a surrogate of LRT bacterial load (r=0.92, p=0.0004).

**Figure 3:**
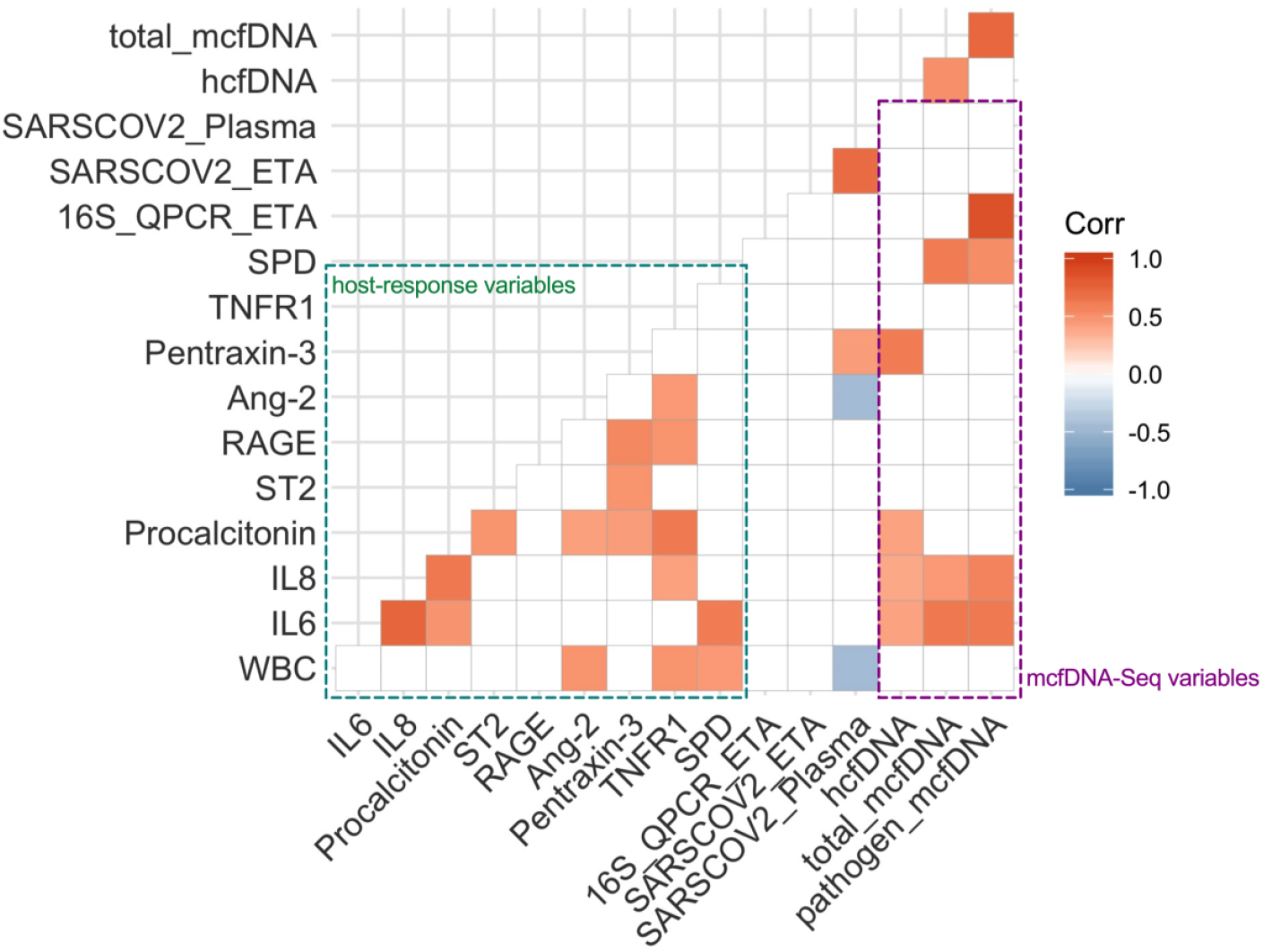
Plasma microbial cell-free DNA levels were significantly correlated with lower respiratory tract bacterial load and plasma host response biomarkers. Correlogram demonstrating comparisons of host nine response biomarkers (green dashed box), number of 16S rRNA gene copies by qPCR in Endotracheal Aspirates (ETA, surrogate for lower respiratory tract bacterial load), number of SARS-COV-2 RNA copies in ETA and plasma samples by qPCR, and mcfDNA-Seq output (hcfDNA, total mcfDNA and pathogen mcfDNA – purple dashed box). Significant correlations with the Pearson’s method and following adjustment for multiple comparisons by the Benjamini-Hochberg method are shown, with direction and strength of correlation depicted by the color scale on the right panel.

### Baseline mcfDNA levels are predictive of 90-day survival

Baseline hcfDNA and total mcfDNA levels were not significantly associated with cumulative mortality at 90 days from ICU admission (Figure 4A-B). However, baseline total mcfDNA levels were significantly associated with worse 90-day survival in a Cox proportional hazards model adjusted for age (hazard ratio for log_10_-transformed mcfDNA 1.30, [1.02-1.64], p=0.03). Stratified by baseline total mcfDNA tertiles (high>1499, middle 344-1499, low<344 MPMs), subjects in the high mcfDNA tertile had worse 90-day survival by Kaplan-Meier curve analysis compared to subjects in other tertiles (global log-rank p=0.02), whereas hcfDNA tertiles were not predictive of 90-day survival (Figure 4C-D). In contrast to the significant prognostic information of baseline total mcfDNA levels, there were no 90-day survival differences between the clinical classification SI groups (log-rank p=0.62 for Micro-SI, Clinical-SI and No-Suspected-SI, data not shown). Stratified by 90-day survival, we found no significant differences in the longitudinal trajectories of total mcfDNA and hcfDNA levels between survivors and non-survivors (e-Figure 5A, B).

**Figure 4:**
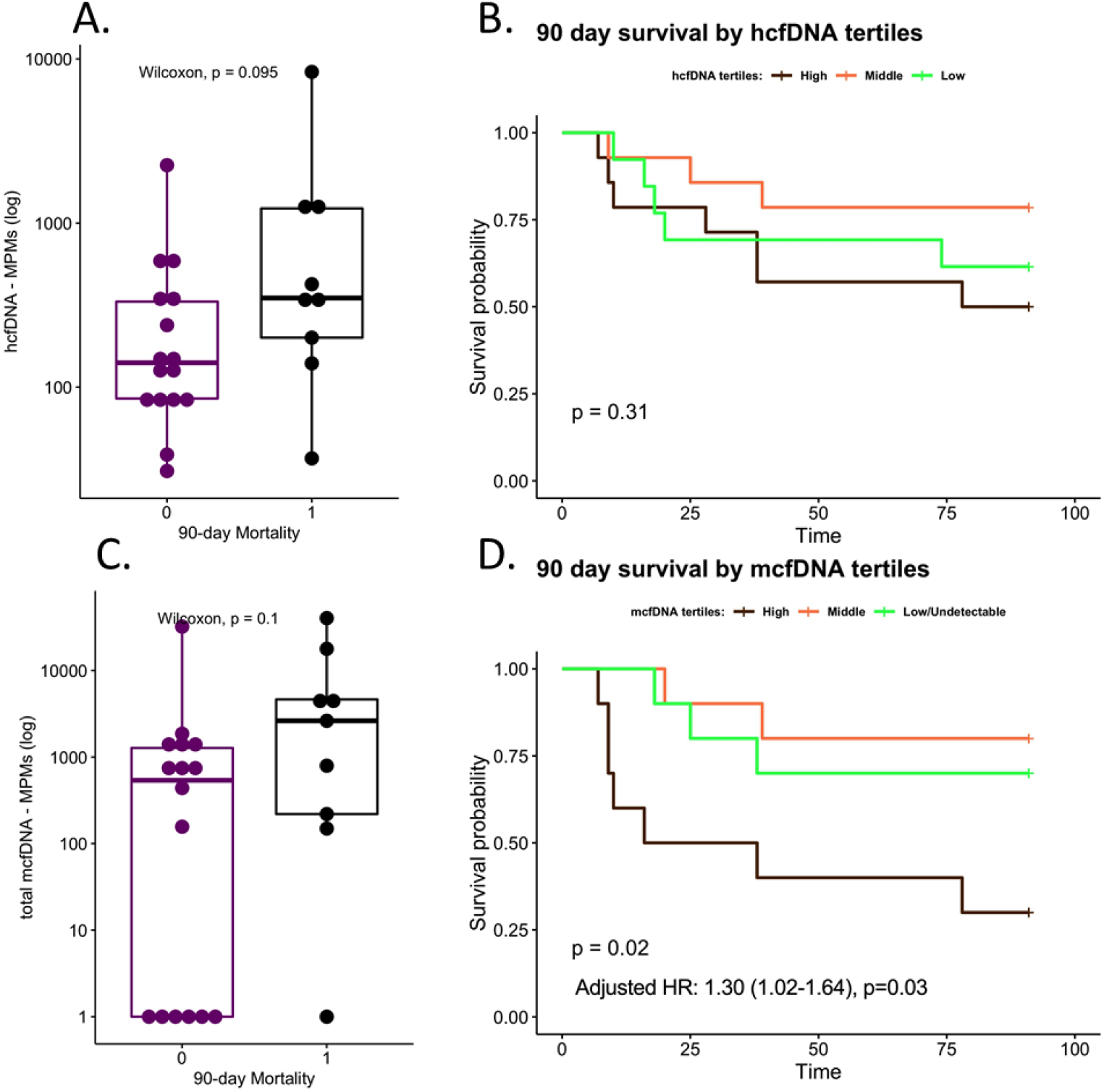
Highest microbial cell-free DNA levels at baseline were significantly associated with worse 90-day survival. 90-day non-survivors had numerically higher (but statistically non-significant) plasma hcfDNA (A) and mcfDNA (C) levels. HcfDNA levels were not significantly associated with 90-day survival by Kaplan-Meier analysis (B). Patients with the highest tertile of mcfDNA (>1499 Molecules per Microliter) had significantly worse survival compared to patients in the other two mcfDNA tertiles (D). In a Cox proportional hazards model adjusted for age, mcfDNA levels were significantly associated with increased hazards of death (D, adjusted hazard ratio for log_10_-transformed mcfDNA 1.30, 95% confidence interval 1.02-1.64, p=0.03).

## Discussion

In this exploratory study, we systematically evaluated 42 COVID-19 inpatients with culture-independent mcfDNA-Seq. We found no significant differences in mcfDNA levels by SI clinical categories at baseline, although culture-confirmed cases of SI (Micro-SI) had higher mcfDNA levels on follow-up samples. The mcfDNA species corresponded to clinically isolated pathogens in 81% of Micro-SI samples, and indicated potentially missed, super-infecting pathogens in up to 41% of subjects for whom there was no clinical suspicion for SI (No-Suspected-SI). Our molecular analyses revealed significant correlations of circulating mcfDNA levels with LRT bacterial burden, suggestive of plausible secondary pneumonias, as well as with plasma host-response biomarkers. Subjects with the highest tertile of mcfDNA burden had significantly lower 90-day survival independent of SI group. Longitudinal assessments revealed that mcfDNA levels can persist in SI without adequate source control such as the patient with parotitis or potentially with antibiotic resistant pathogens such as the patient with resistant *Pseudomonas* VAP. Our results were also notable for an unexpectedly high rate of failed mcfDNA-Seq runs due to high burden of circulating hcfDNA, which was much higher than historic cases of non-COVID-19 pneumonia, perhaps due to significant human tissue damage in severely ill patients with COVID-19, including patients supported by extra-corporeal membrane oxygenation.

The higher likelihood of subsequent failed samples after initial failed samples suggests subject-specific influences on sequencing run outputs and results.

Our study highlights the challenge of clinically distinguishing cases with isolated SARS-CoV-2 infection from those complicated by SIs. The results of host-response biomarker analyses and mcfDNA-Seq did not match the clinical SI diagnoses and clinically-directed microbiologic work-up. For example, procalcitonin levels are often used as diagnostic biomarkers for bacterial infections, but have unclear diagnostic value for SIs in COVID-19.^20,21^ We found similar procalcitonin levels among clinical SI groups, and importantly no significant association with mcfDNA levels. The clinical validity of mcfDNA sequencing was further supported by the significant correlations with LRT bacterial burden and systemic host-response biomarkers, as well as by the predictive value on 90-day survival. Our study suggests that noninvasive mcfDNA sequencing may offer insights into the presence of an SI and its impact on outcome.

Circulating mcfDNA was significantly associated with host immune response in COVID-19. McfDNA species may reflect SI-causal pathogens (regardless of their viability in the bloodstream), but may also signify lung barrier disruption in patients with acute lung injury from COVID-19, or even gut barrier disruption in patients with circulatory shock and hypoperfusion.^14^ Thus, positive mcfDNA calls should not be directly interpreted as evidence of superinfecting pathogens, but interpreted within the context of a critical illness with colonized mucosal surfaces by microbiota and impaired barrier function. The significant correlation between hcfDNA levels and RALE scores suggests that lung injury may account, at least in part, for the systemic levels of circulating hfcDNA. The significant correlations of mcfDNA with IL-6 and IL-8 levels highlighted the potential importance of mcfDNA in the inflammatory cascade of COVID-19, since mcfDNA can be recognized by Toll-like receptors and propagate systemic inflammatory responses, independent of the effects of SARS-CoV-2 on innate immunity.^22^ Additionally, the case of Herpesvirus DNA identification highlighted the prevalence of viral re-activation in critical illness. Thus, any positive mcfDNA signal requires careful interpretation, as it may represent superinfecting bacterial or fungal VAP pathogens, translocating commensal microbes, sample collection skin/environmental contaminants or re-activated viral organisms.

Our study has several limitations. The sample size of 42 subjects limits the statistical power for some of the analyses, yet it is to our knowledge the largest study to utilize plasma metagenomics in patients with COVID-19, and we found significant associations with host inflammation and outcomes, consistent with our hypotheses. Clinical samples for microbiologic workup were collected at the discretion of the treating clinicians, and thus some of the mcfDNA-culture comparisons are limited. Nonetheless, our clinical dataset is representative of standard of care at a tertiary academic center. We systematically examined all patients for molecular evidence of possible SI using systematic evaluation with mcfDNA testing, regardless of clinician impressions. It is possible that for certain No-Suspected-SI subjects, the mcfDNA reported may not represent an active infection. Careful review of the mcfDNA-Seq output and integration with available evidence on organismal pathogenicity can help further inform interpretation of pathogen mcfDNA. Nonetheless, circulating mcfDNA carries prognostic information underlined by the significant associations with systemic inflammation and survival.^11^ Our study was limited by an unexpected amount of failed mcfDNA-Seq analyses due to high amounts of interfering hcfDNA, but this finding provided important insight into the high degree of human cellular damage in COVID-19.

Our study advances understanding of the pathobiologic and diagnostic importance of circulating mcfDNA in COVID-19 and demonstrates limitations in clinical assessment of SIs. McfDNA correlated with biomarkers of host immune response and LRT microbial burden. Additional characterization of host inflammatory response components and triggers in COVID-19 is essential in the era of immunomodulatory therapeutics. The noninvasive modality of quantifying mcfDNA load and accurately identifying microbial species may offer a sensitive tool for SI detection, as well as further advance our understanding of the role of translocating microbiota in critical illness. McfDNA sequencing may be particularly helpful in spontaneously breathing patients on high flow nasal cannula or non-invasive mechanical ventilation, for whom access to LRT specimens is challenging. Further prospective investigation for treatment guidance is necessary to systematically evaluate the incidence of molecular evidence for SI and examine the impact of non-invasive screening on patient outcomes.

## Data Availability

All data produced in the present work are contained in the manuscript

https://pubmed.ncbi.nlm.nih.gov/33888575/

## SUPPLEMENT

**e-Table 1:** Baseline characteristics by sequencing run success.

| Variable | Pass | Pass Qualitatively | Fail | P-value |
| --- | --- | --- | --- | --- |
| N | 25 | 5 | 12 |  |
| Male, n (%) | 20 (64.5) | 1 (20.0) | 8 (61.5) | 0.12 |
| Age (median [IQR]) | 68.7 [60.2, 76.8] | 78.4 [63.1, 86.8] | 58.5 [57.2, 64.7] | **0.04** |
| BMI (median [IQR]) | 31.6 [26.4, 34.0] | 30.1 [26.8, 36.4] | 39.6 [28.7, 42.4] | 0.35 |
| Diabetes, n (%) | 11 ( 44.0) | 3 ( 60.0) | 4 ( 33.3) | 0.59 |
| COPD, n (%) | 9 ( 36.0) | 1 ( 20.0) | 0 ( 0.0) | **0.05** |
| Immunosuppression, n (%) | 2 ( 8.0) | 1 ( 20.0) | 1 ( 8.3) | 0.7 |
| Invasive Mechanical Ventilation, n (%) | 14 ( 56.0) | 4 ( 80.0) | 9 ( 75.0) | 0.39 |
| ECMO, n (%) | 3 ( 12.0) | 1 ( 20.0) | 4 ( 33.3) | 0.3 |
| WBC (median [IQR]) | 9.4 [6.3, 11.8] | 11.1 [11.1, 11.5] | 10.2 [7.4, 13.7] | 0.46 |
| WHO ordinal scale at ICU admission (median [IQR]) | 5.0 [5.0, 6.0] | 5.5 [5.0, 6.5] | 7.0 [6.0, 8.5] | **0.01** |
| RALE score (median [IQR]) | 22.5 [21.0, 28.0] | 23.0 [22.8, 24.0] | 27.5 [20.6, 35.8] | 0.56 |
| SARS-CoV-2 RNA in Plasma (cps/mL, median [IQR]) | 196.5 [3.0, 3349.0] | 6352.0 [3286.0, 26029.3] | 2491.0 [335.0, 6874.5] | 0.3 |
| IL6 pg/ml (median [IQR]) | 33.9 [16.9, 124.9] | 165.7 [89.7, 1109.9] | 51.1 [15.7, 221.3] | 0.19 |
| IL8 pg/ml (median [IQR]) | 22.2 [13.9, 33.6] | 35.9 [29.9, 181.2] | 17.6 [15.1, 62.3] | 0.29 |
| ST2 pg/ml (median [IQR]) | 142138.6 [78170.4, 198277.7] | 253415.2 [218252.9, 578677.0] | 287500.3 [216718.3, 328329.0] | **<0.01** |
| TNFR1 pg/ml (median [IQR]) | 4186.0 [3555.3, 7971.9] | 9493.9 [6465.9, 12598.6] | 4919.9 [3417.1, 6956.2] | 0.44 |
| SPD pg/ml (median [IQR]) | 24.3 [10.0, 43.0] | 80.5 [20.8, 141.9] | 26.0 [13.2, 53.1] | 0.46 |
| RAGE pg/ml (median [IQR]) | 4245.9 [2419.5, 9023.2] | 4518.6 [2410.7, 9982.9] | 5518.4 [2884.1, 7936.1] | 0.97 |
| Ang-2 pg/ml (median [IQR]) | 3905.8 [1935.6, 7130.4] | 4505.1 [3942.7, 8903.3] | 4543.4 [2886.8, 10443.9] | 0.5 |
| Procalcitonin pg/ml (median [IQR]) | 280.0 [167.0, 558.5] | 3630.8 [1767.8, 4556.7] | 580.8 [240.5, 1478.0] | **0.01** |
| Pentraxin-3 pg/ml (median [IQR]) | 6085.9 [2937.8, 13127.0] | 19508.6 [6766.4, 32196.3] | 12979.7 [7122.9, 18778.3] | 0.1 |
| Human cfDNA MPM (median [IQR]) | 200.3 [91.3, 424.2] | 722.2 [686.2, 1299.7] | 1343.1 [949.5, 3731.9] | **<0.01** |
| **Total microbial cfDNA MPM (median [IQR])** | 794.8 [1.0, 1863.1] | 1480.8 [818.1, 21805.9] | NA | 0.14 |
| **Bacterial cfDNA MPM (median [IQR])** | 252.3 [1.0, 1343.9] | 1480.8 [537.1, 21805.9] | 42735.5 [42735.5, 42735.5] | 0.13 |
| **Fungal cfDNA MPM (median [IQR])** | 1.0 [1.0, 1.0] | 1.0 [1.0, 1.0] | 1.0 [1.0, 1.0] | 0.67 |
| **Respiratory pathogen cfDNA MPM (median [IQR])** | 1.0 [1.0, 1015.8] | 1480.8 [1.0, 21805.9] | 42735.5 [42735.5, 42735.5] | 0.12 |

P-values significant below threshold of 0.05 are shown in bold. Abbreviations: Microbiologically-Diagnosed Secondary Infection (Micro-SI); Clinically-Diagnosed secondary infections (Clinical-SI); No Clinical Suspicion for SI (No-Suspected-SI); chronic obstructive pulmonary disease (COPD); extracorporeal membrane oxygenation (ECMO); Radiographic Assessment of Lung Edema score (RALE score); receptor for advanced glycation end products (RAGE); suppression of tumorigenicity (ST-2); tumour necrosis factor receptor (TNFR-1); Surfactant Protein D (SPD); molecules per microliter (MPMs).

**e-Figure 1:**
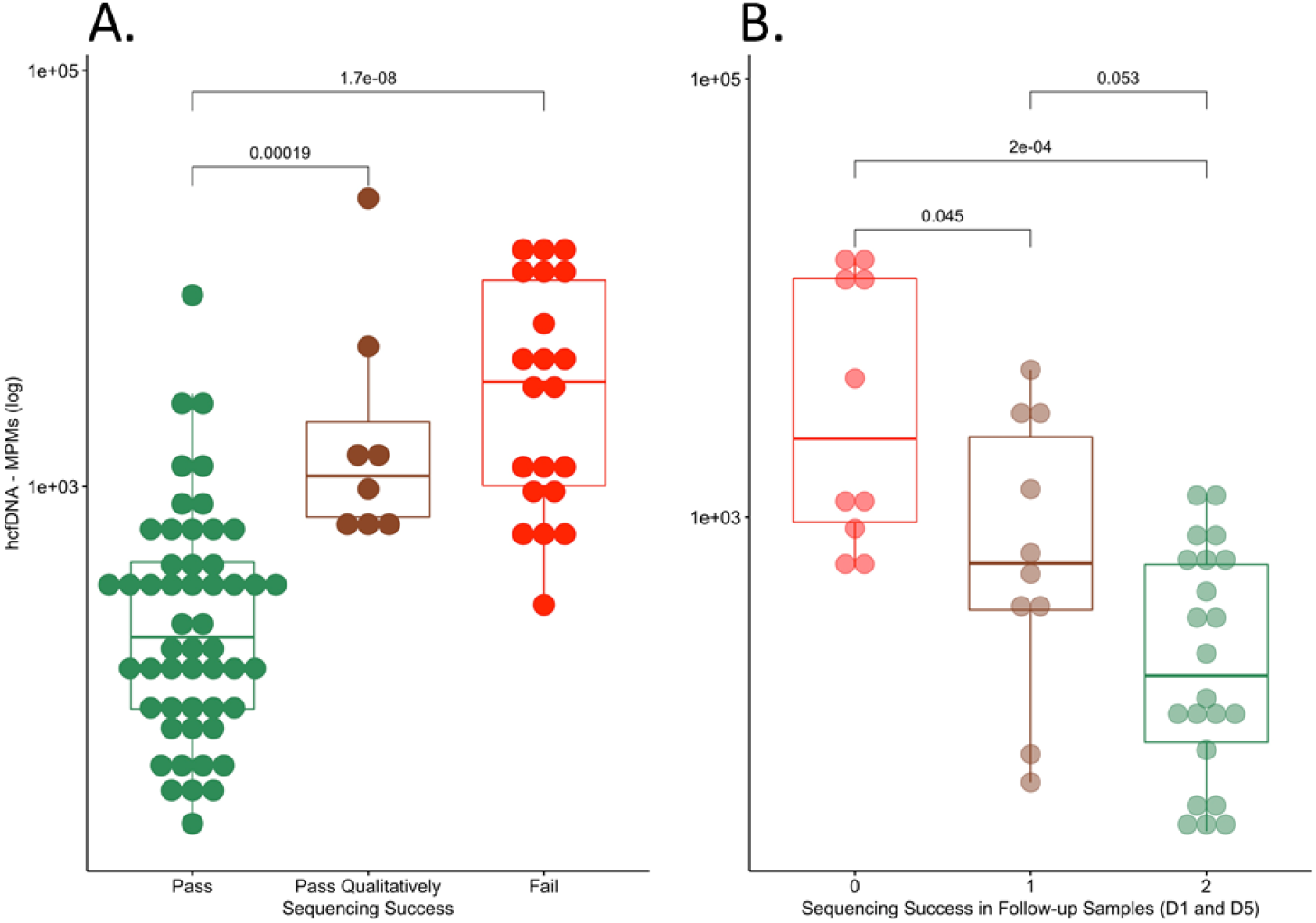
Successful plasma metagenomic sequencing runs had significantly lower levels of human cell-free DNA compared to unsuccessful runs. We classified the derived metagenomic sequences as human (hcfDNA) vs. microbial (mcfDNA), expressed as molecules per microliter (MPMs). Based on meeting minimum sequencing coverage metric required for quality control, we classified sequencing runs as successful (“Pass”), “Qualitatively Pass” or “Failed”. Baseline “Pass” samples had significantly lower hcfDNA compared to “Qualitatively Pass” or “Failed” samples (Wilcoxon test pairs p<0.001, panel A). We also found that among subjects with both Day 1 and Day 5 samples, those samples that failed on both time points per subject (i.e. “0” sequencing success in panel B) had significantly higher hcfDNA levels compared to “Pass” samples on both days (Wilcoxon p-value <0.001).

**e-Figure 2:**
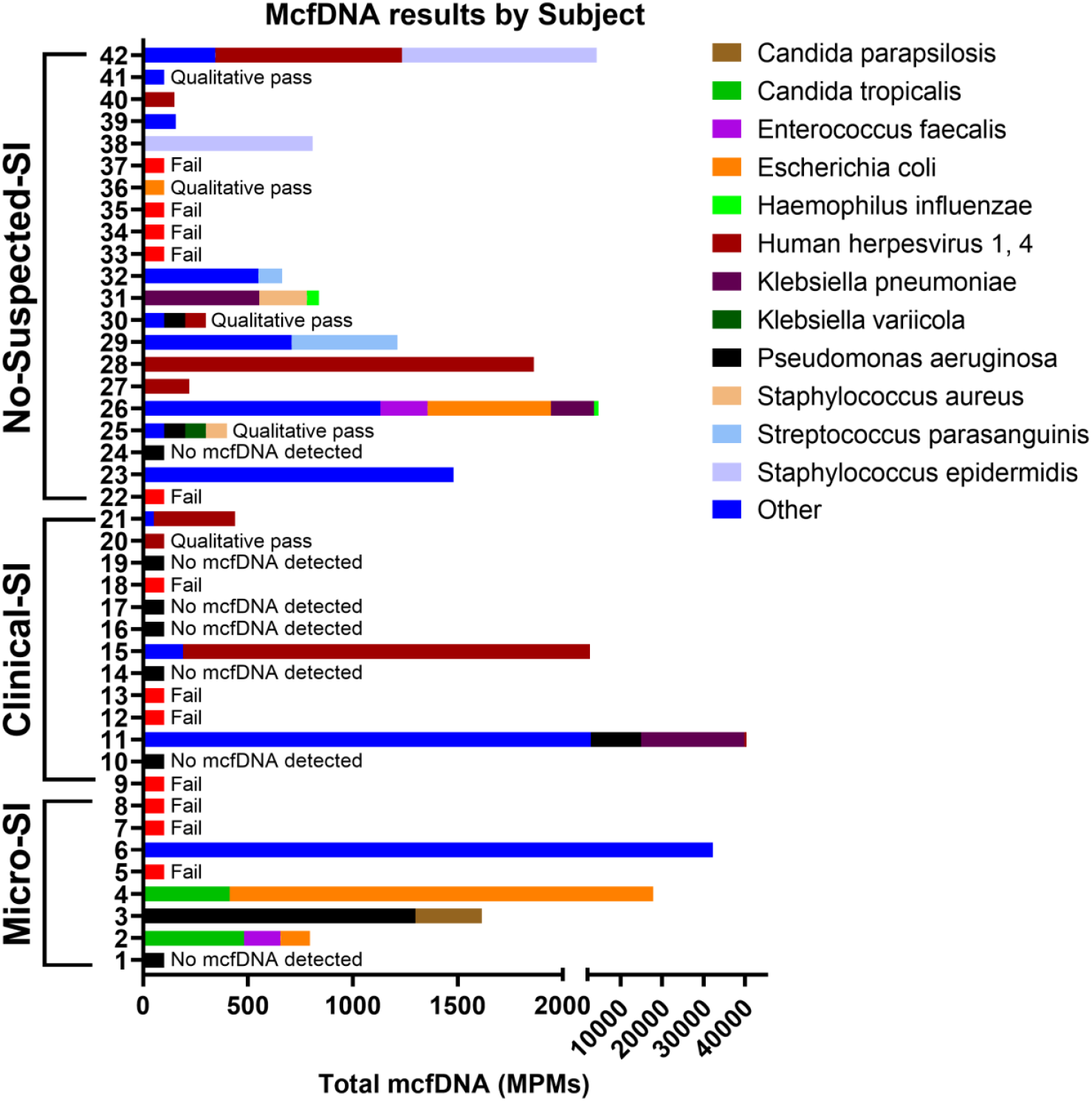
Significantly higher proportion of positive samples with mcfDNA species calls in No-Suspected-SI compared to Clinical-SI samples. We display samples grouped by SI classification and sample identifier, showing the species and respective MPMs called by mcfDNA-Seq. Standardized bars indicate failed samples, samples with no species called, and those with qualitative pass, yielding species each denoted with standardized bar. Infrequently called species were combined for visual simplicity.

**e-Figure 3:**
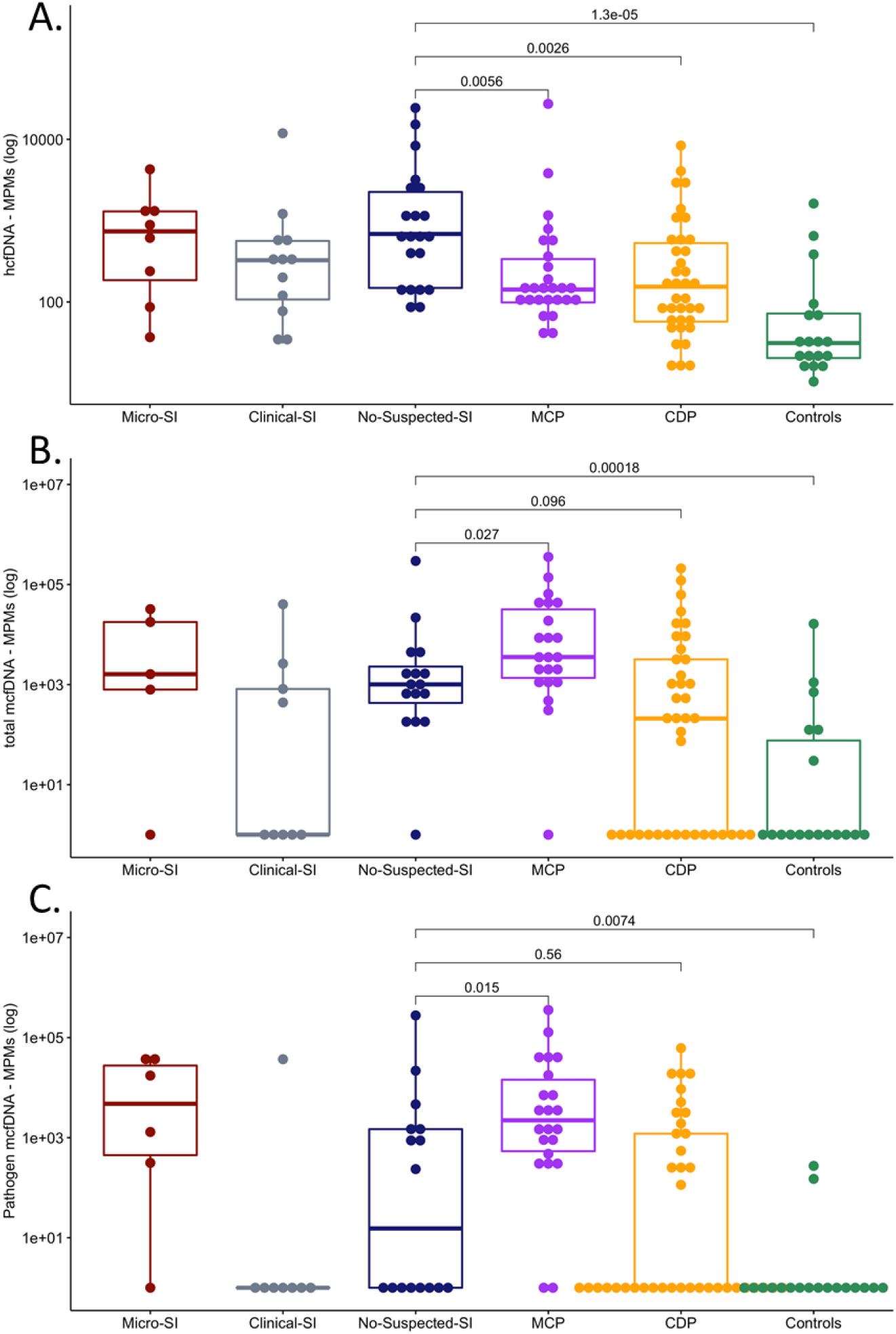
Subjects with COVID-19 have much higher levels of human cell-free DNA compared to non-COVID subjects with and without pneumonia. To contextualize the circulating cfDNA load in our COVID-19 cohort, we compared hcfDNA, total mcfDNA and pathogen mcfDNA MPMs between the COVID-19 SI categories against available published data from our group for mechanically ventilated patients with microbiologically-confirmed pneumonia (n=26, MCP), clinically-diagnosed pneumonia (n=41, CDP) and uninfected controls (n=16, intubated for airway protection or due to cardiogenic pulmonary edema). We found markedly higher levels of hcfDNA in subjects with COVID-19 compared to all non-COVID patient groups (p-values shown for the No-Suspected-SI only for parsimony). Non-COVID patients with microbiologically-confirmed pneumonia had higher mcfDNA levels compared to patients with COVID-19 with No-Suspected-SI, who in turn had markedly higher mcfDNA levels compared to uninfected controls.

**e-Figure 4:**
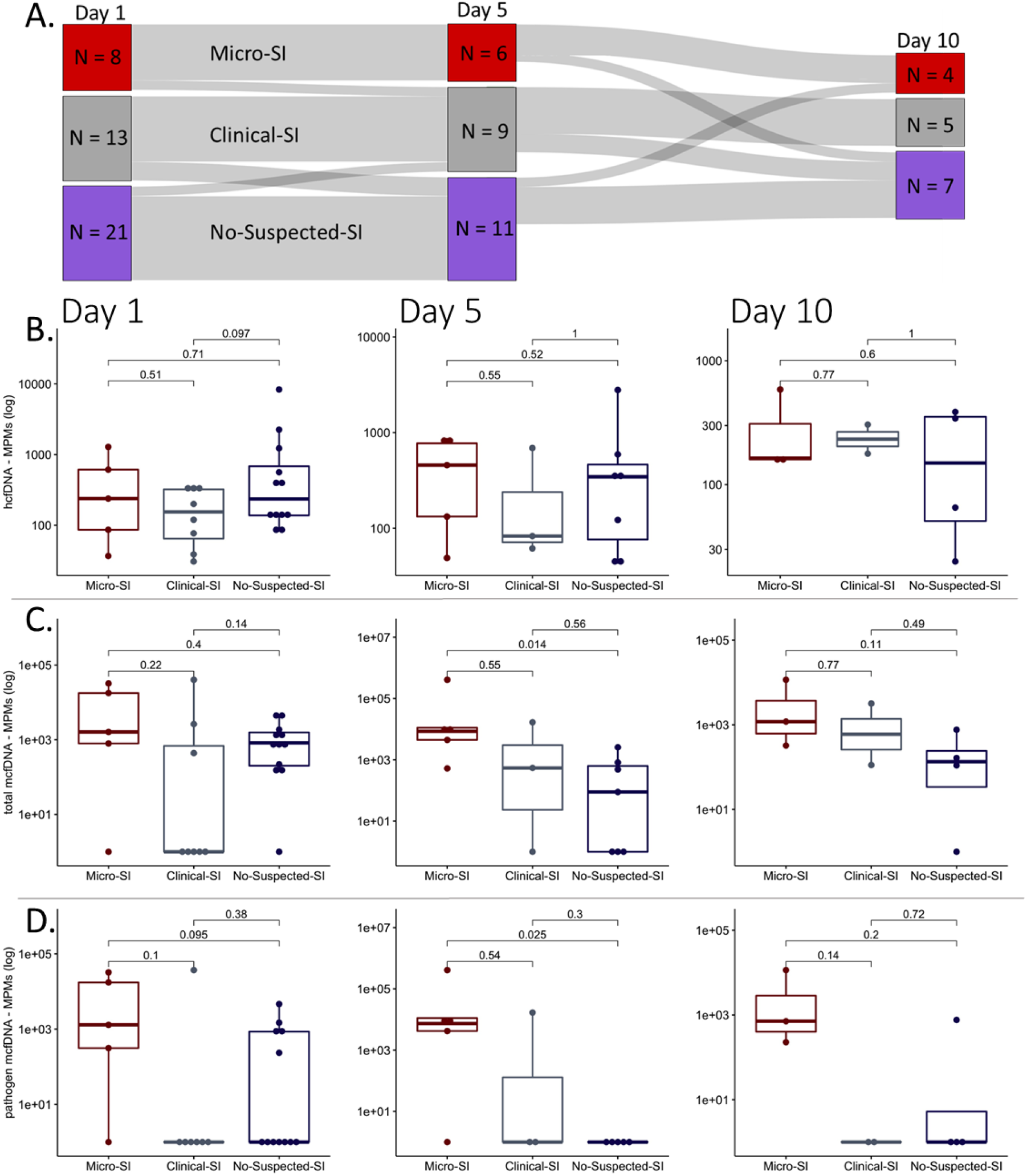
Longitudinal subject, mcfDNA and human cfDNA changes across SI classification. This Sankey plot shows the subjects who transitioned between SI categories throughout the study from days 1, to 5, and 10 (A). Height of bars represents number of subjects. Attrition occurred throughout the study leading to decreased height of day 5 and 10 nodes. HcfDNA was not significantly different among SI groups across the study period (B). Total mcfDNA was not significantly different among SI categories on enrollment (C). Micro-SI subjects had significantly higher total and pathogen mcfDNA versus No-Suspected-SI subjects at Day 5 (C, D).

**e-Figure 5:**
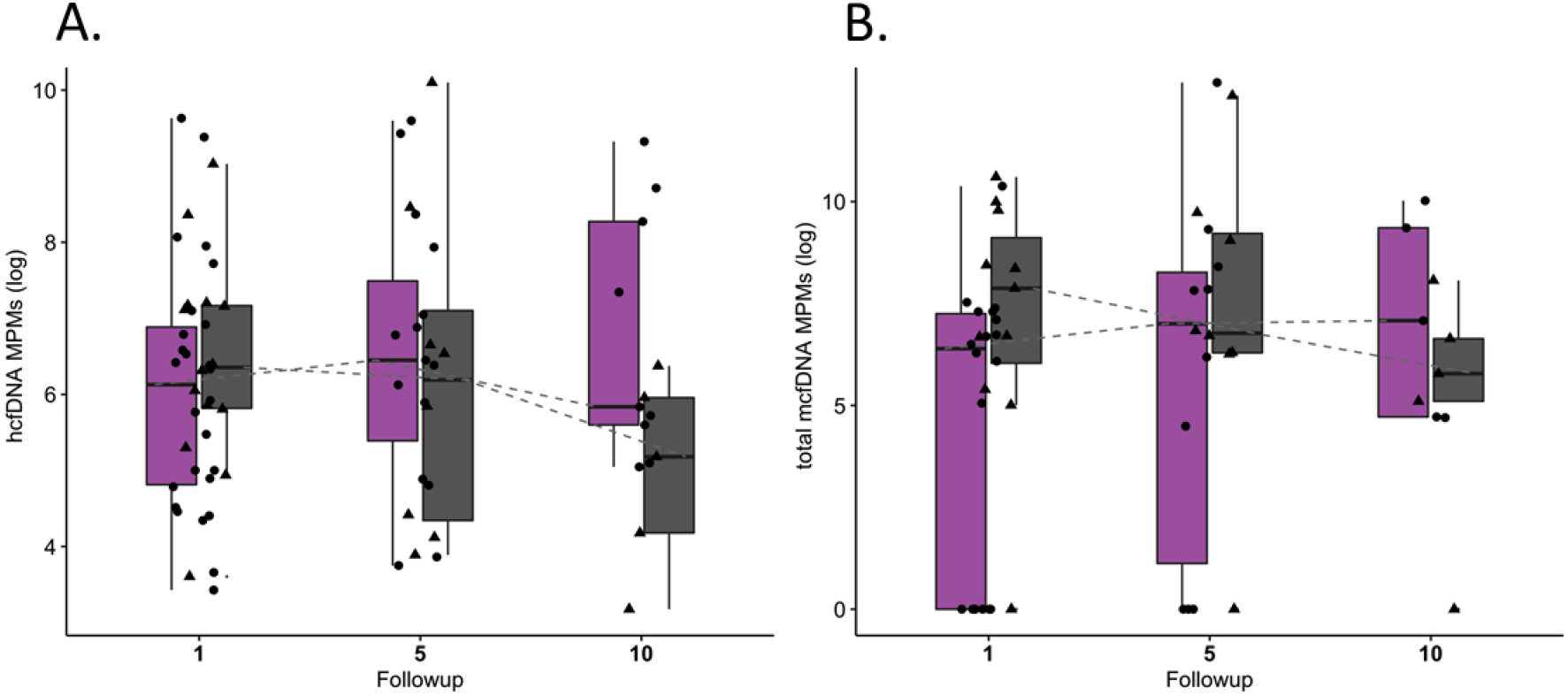
Human and total mcfDNA trajectories among survivors and non-survivors. Trajectories of human cfDNA and total mcfDNA were not significantly different across the study period in survivors and non-survivors (A, B).

**e-Table 2:**
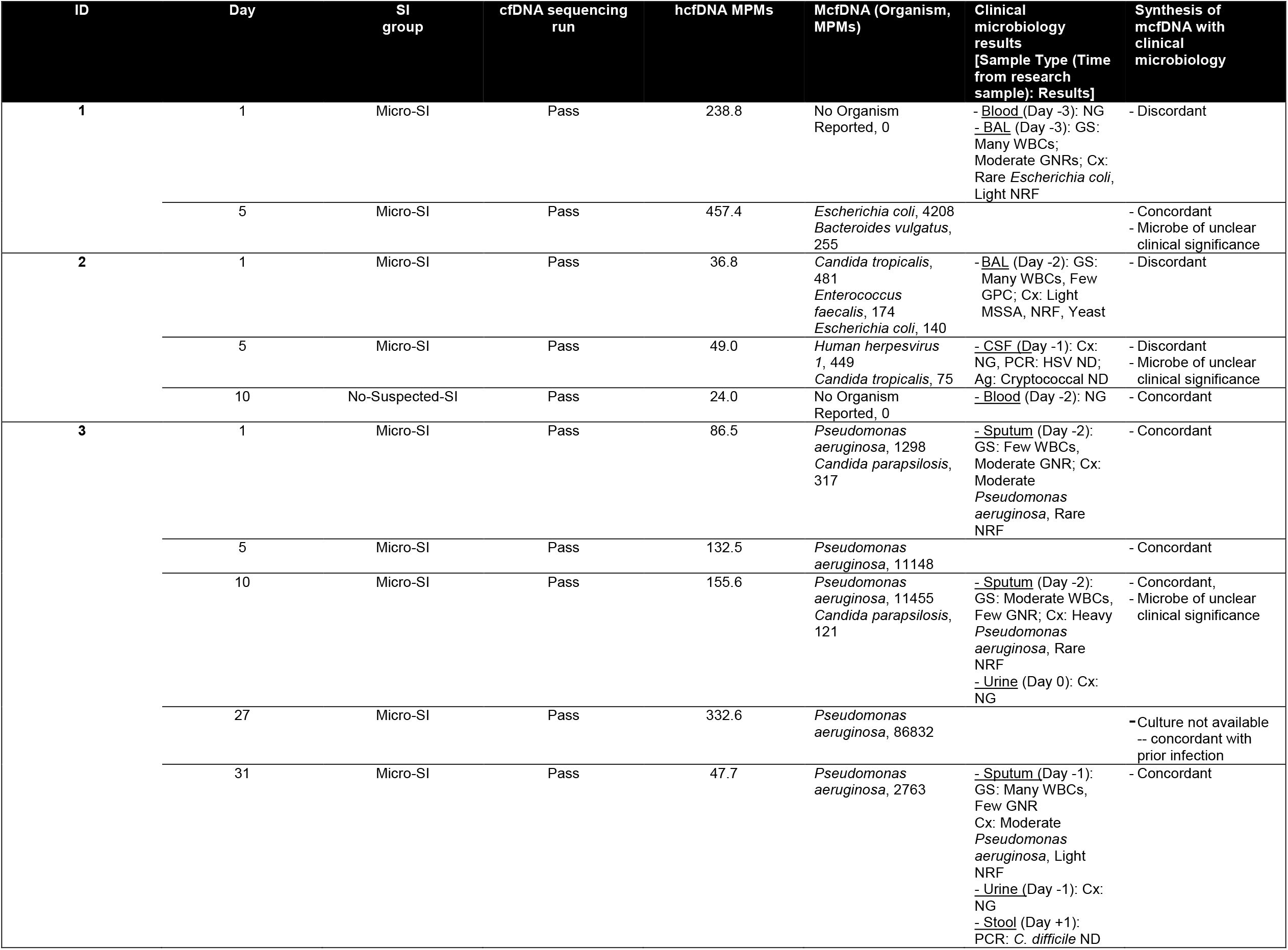
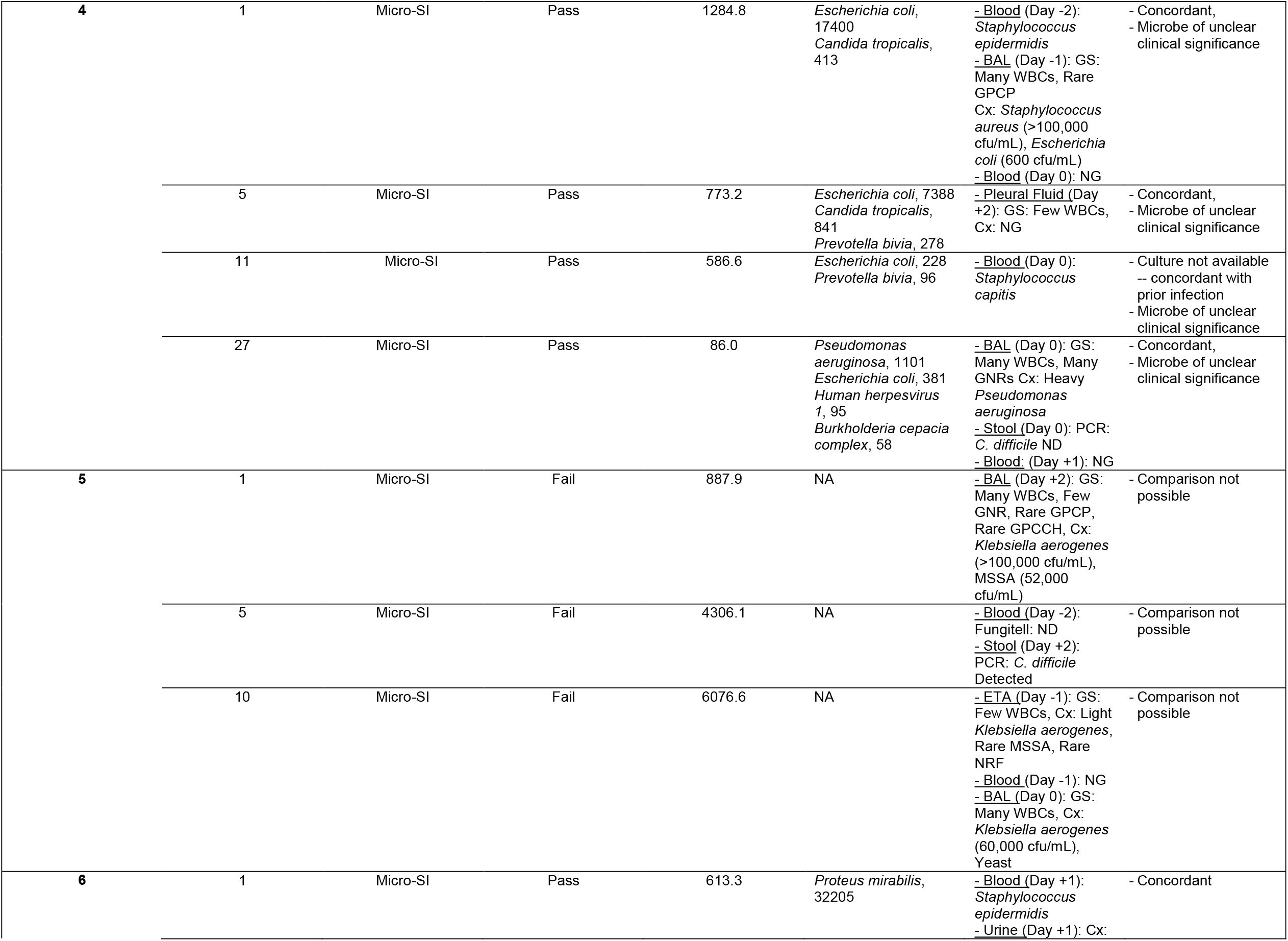
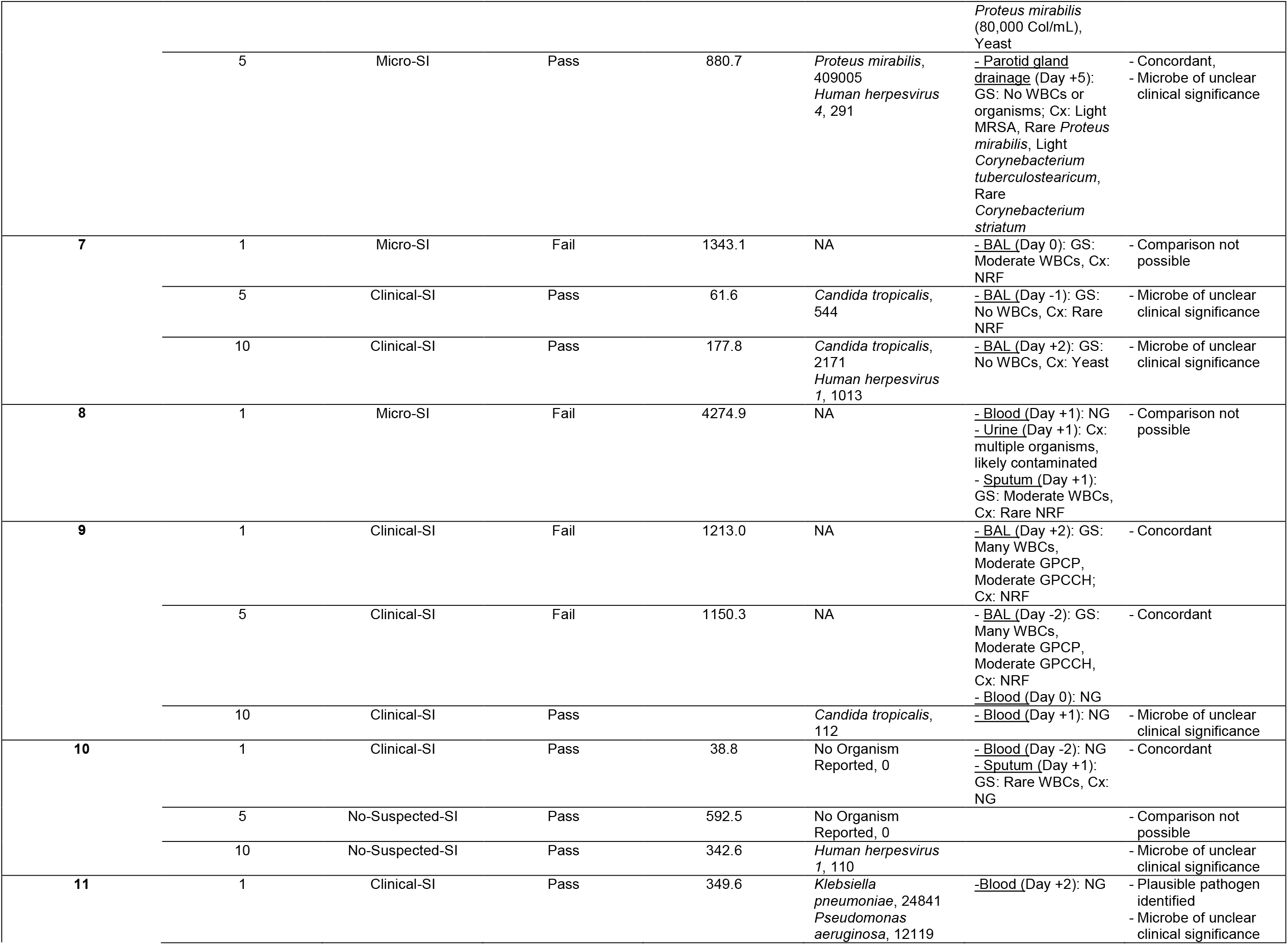
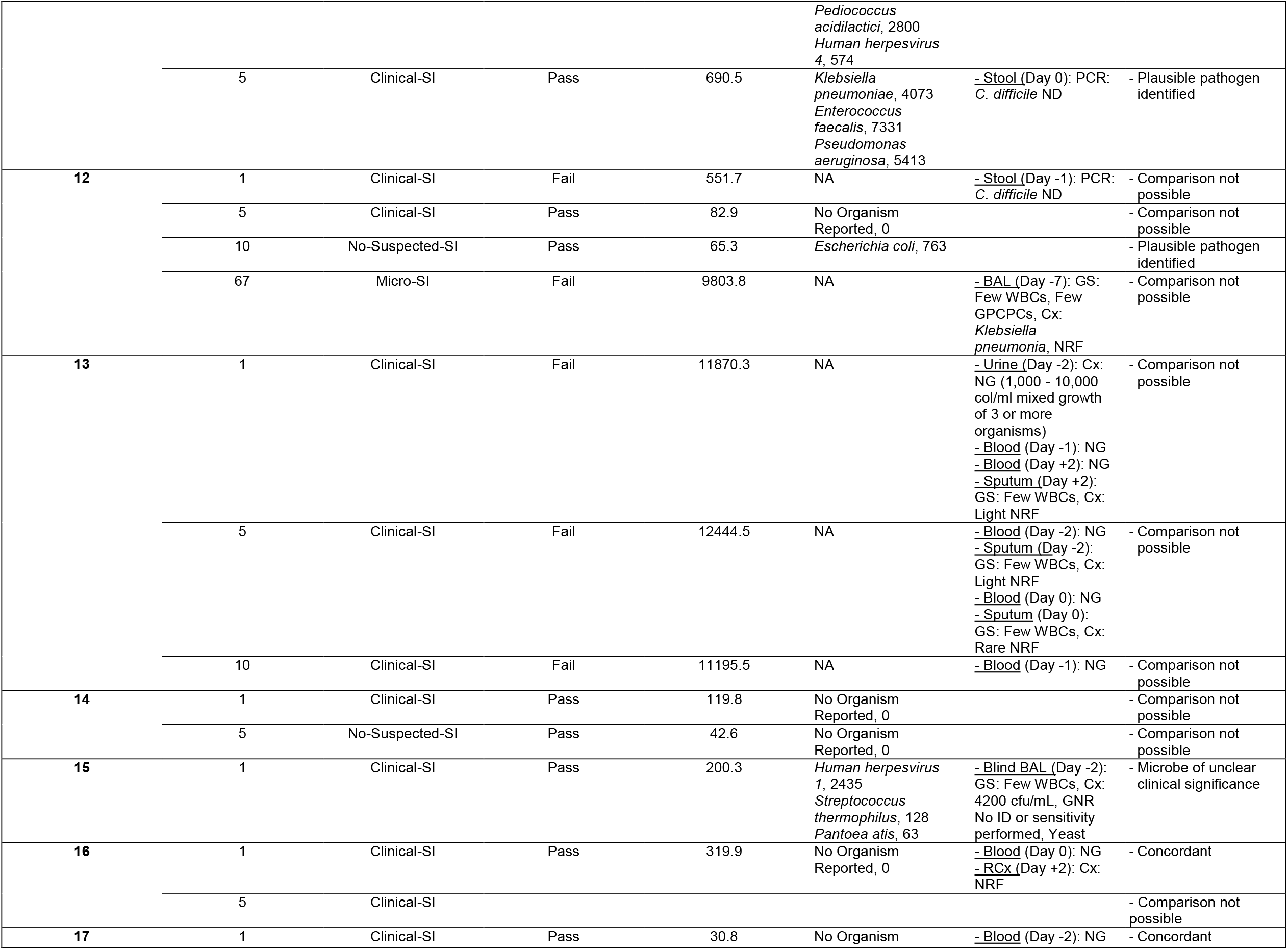
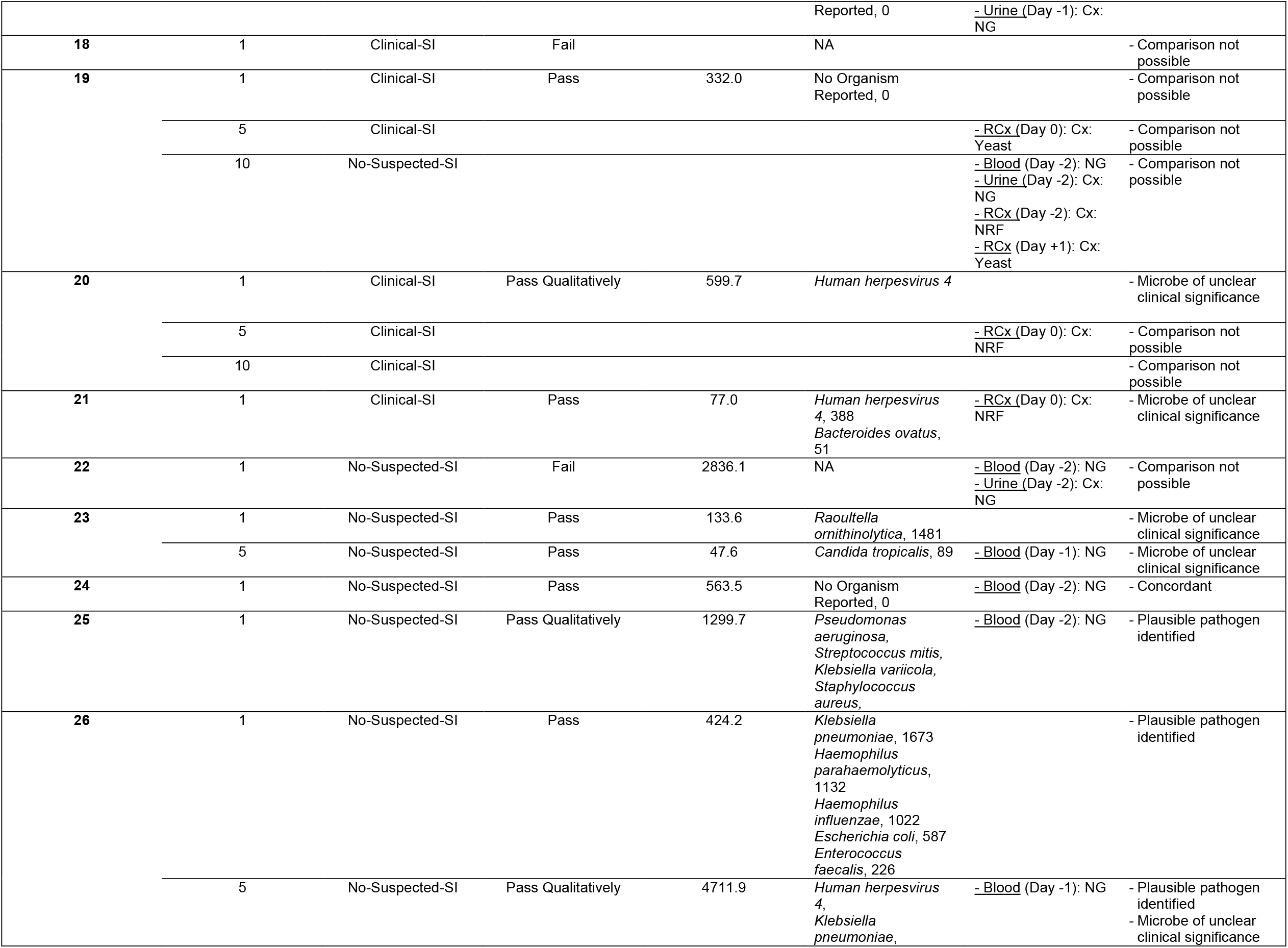
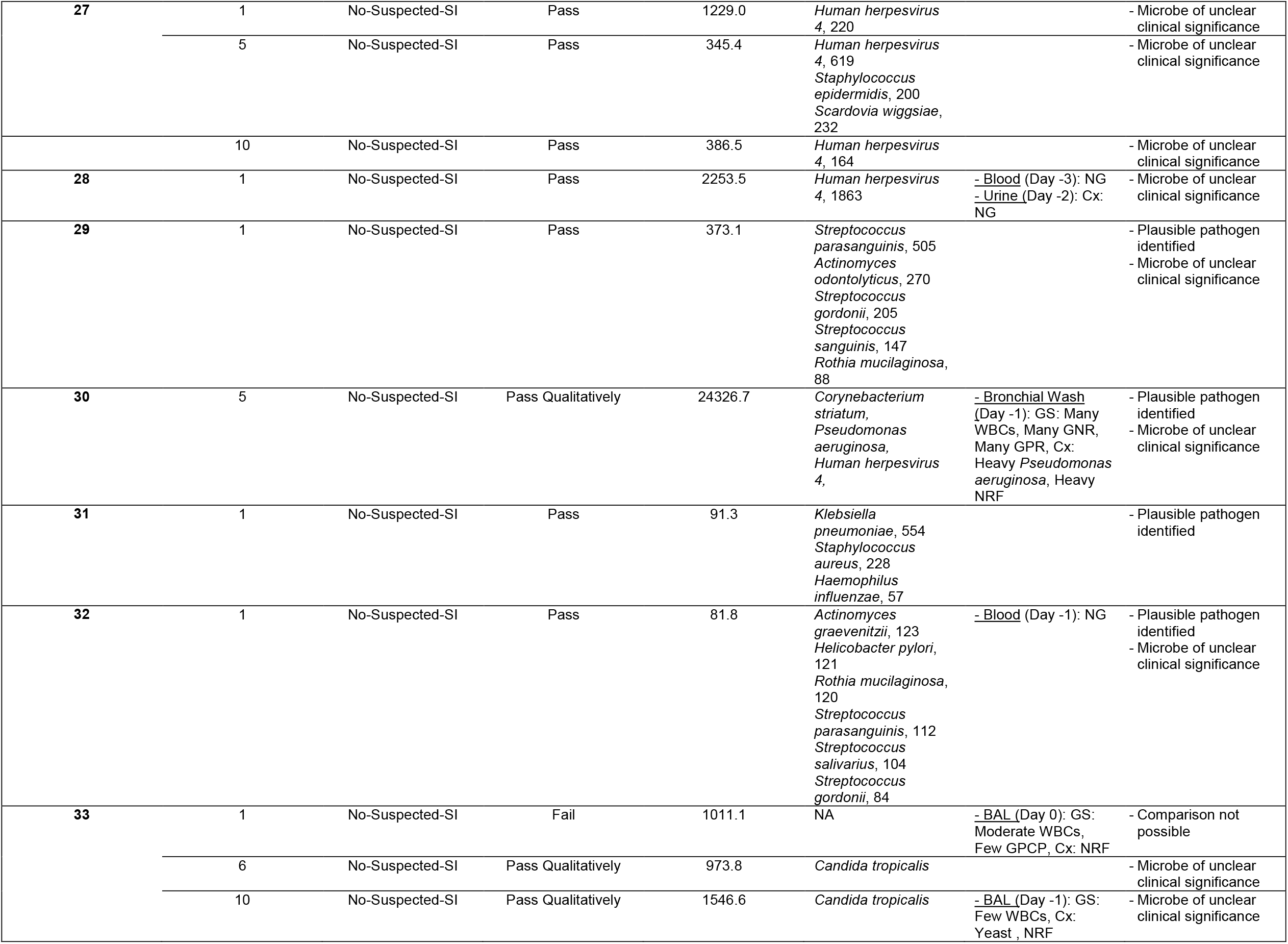
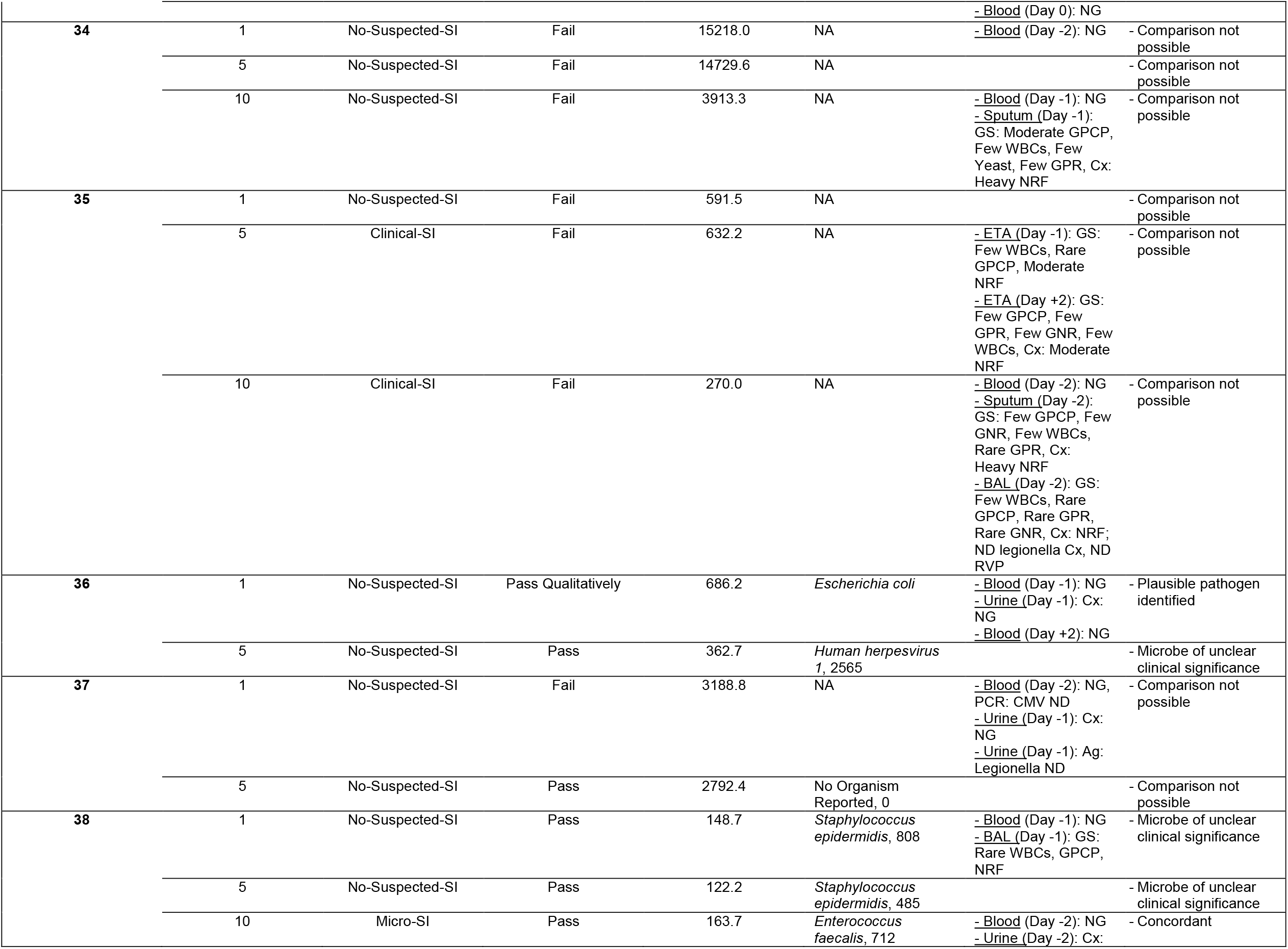
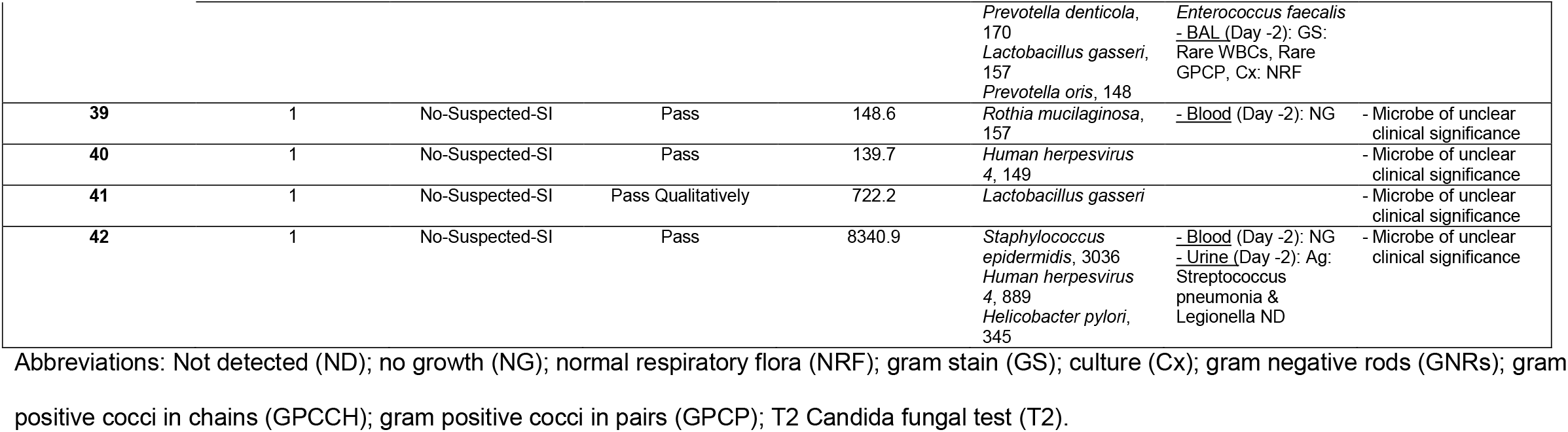
Subject cfDNA and culture results.

**e-Table 3:** Microbial species classifications.

| Taxon | Class | Anatomic Compartment |
| --- | --- | --- |
| *Actinomyces graevenitzi* | Recognized pathogen | Oral |
| *Actinomyces odontolyticus* | Recognized pathogen | Oral |
| *Bacteroides ovatus* | Microbe with unclear clinical importance | GI |
| *Bacteroides vulgatus* | Microbe with unclear clinical importance | GI |
| *Burkholderia cepacia complex* | Recognized pathogen | Respiratory, Nosocomial |
| *Candida parapsilosis* | Microbe with unclear clinical importance | Skin, GI |
| *Candida tropicalis* | Microbe with unclear clinical importance | Skin, GI |
| *Corynebacterium striatum* | Microbe with unclear clinical importance | Skin |
| *Enterococcus faecalis* | Recognized pathogen | GI |
| *Enterococcus faecium* | Recognized pathogen | GI |
| *Escherichia coli* | Recognized pathogen | GI |
| *Haemophilus influenzae* | Recognized pathogen | Oral |
| *Haemophilus parahaemolyticus* | Microbe with unclear clinical importance | Oral |
| *Helicobacter pylori* | Microbe with unclear clinical importance | GI |
| *Human herpesvirus 1* | Microbe with unclear clinical importance | Unknown |
| *Human herpesvirus 4* | Microbe with unclear clinical importance | Unknown |
| ***Klebsiella pneumoniae*** | Recognized pathogen | Oral, GI |
| ***Klebsiella variicola*** | Recognized pathogen | GI |
| ***Lactobacillus gasseri*** | Microbe with unclear clinical importance | GI |
| ***Pantoea ananatis*** | Microbe with unclear clinical importance | Unknown |
| ***Pediococcus acidilactici*** | Microbe with unclear clinical importance | GI |
| ***Prevotella bivia*** | Microbe with unclear clinical importance | Oral |
| ***Prevotella denticola*** | Microbe with unclear clinical importance | Oral |
| ***Prevotella oris*** | Microbe with unclear clinical importance | Oral |
| ***Proteus mirabilis*** | Recognized pathogen | GI |
| ***Pseudomonas aeruginosa*** | Recognized pathogen | GI, Pulmonary, Nosocomial |
| ***Raoultella ornithinolytica*** | Recognized pathogen | GI |
| ***Rothia mucilaginosa*** | Microbe with unclear clinical importance | Oral |
| ***Staphylococcus aureus*** | Recognized pathogen | Skin |
| ***Staphylococcus epidermidis*** | Microbe with unclear clinical importance | Skin |
| ***Streptococcus gordonii*** | Microbe with unclear clinical importance | Oral |
| ***Streptococcus infantarius*** | Microbe with unclear clinical importance | Oral |
| ***Streptococcus mitis*** | Recognized pathogen | Oral, Skin |
| ***Streptococcus parasanguinis*** | Recognized pathogen | Oral |
| ***Streptococcus salivarius*** | Microbe with unclear clinical importance | Oral |
| ***Streptococcus sanguinis*** | Microbe with unclear clinical importance | Oral |
| ***Streptococcus thermophilus*** | Microbe with unclear clinical importance | Oral |
| ***Scardovia wiggisiae*** | Microbe with unclear clinical importance | Oral |

